# Mapping the evidence on environmental health services in healthcare facilities in low- and middle-income countries: A systematic literature inventory of over 4,000 studies

**DOI:** 10.1101/2025.11.26.25341056

**Authors:** Lucy Tantum, Darcy M. Anderson, Emily P. Jones, Ryan Cronk

## Abstract

**Background:** Environmental health services in healthcare facilities —including water, sanitation, hygiene, waste management, cleaning, and infection control—prevent disease and strengthen healthcare delivery. Yet environmental health service provision is inadequate in many low- and middle-income countries (LMICs). Despite the importance of monitoring and improving services, no comprehensive evidence map exists to describe global knowledge and gaps for action and improvement. The study objectives were to comprehensively catalog published literature on environmental health services in healthcare facilities in LMICs by service domain, study type, and relevance to policy and practice.

**Methods:** We conducted a systematic literature search in 2023 and performed an update in 2025. Through a title/abstract screening and tagging process, we developed a literature inventory that categorized studies by topic, design, and relevance to policy and practice objectives.

**Results:** The literature inventory included 4,381 studies. Fifty-eight percent of the studies were baseline assessments of environmental health services, 36% involved formative research (e.g., qualitative methods), and 13% evaluated interventions or implementation strategies. Most studies (62%) examined hygiene at points of care, while 9% examined water and 6% sanitation. Twenty-seven percent of studies examined services in the context of the COVID-19 pandemic.

**Conclusions:** There is little evidence for effective interventions and implementation strategies to improve and sustain environmental health services, especially for water and sanitation services. Formative research on under-studied services can help policymakers and practitioners identify areas to prioritize investment and programming. Findings can inform the development of research agendas and practical guidelines for improving access to safe healthcare environments.

## 1. Introduction

A critical challenge hampering health systems in low-and middle-income countries (LMICs) is the lack of environmental health services in healthcare facilities, including water, sanitation, hygiene, waste management, cleaning, and laundry (World Health Organization (WHO), 2015). An estimated 22% of healthcare facilities lack basic water service, and 33% lack basic hand hygiene services (Joint Monitoring Programme, 2024). Environmental health services are important to reduce healthcare-associated infection and the development of antimicrobial resistance (Maina et al., 2019; Peters et al., 2022; Watson et al., 2019), support patient and healthcare worker satisfaction and workforce retention (Anderson et al., 2023; Bouzid et al., 2018; Fitzpatrick et al., 2025), prepare for a rapid and effective pandemic response (Howard et al., 2020), and support health systems to reach targets for universal primary care through providing essential infrastructure, equipment, and supplies to enable care delivery (World Health Organization and United Nations Children’s Fund, 2025).

While the importance of environmental health services for safe and effective medical care has been well established for decades—early evidence of the importance of hand hygiene for infection prevention and control dates to the mid-19^th^ century (Pittet and Boyce, 2001; Semmelweis, 1861)—explicit recognition of environmental health services in healthcare facilities as a clear and consolidated target on the global development agenda is relatively recent.

Healthcare facilities were first recognized as a new priority setting for environmental health services in 2015 under the Sustainable Development Goals. Goal 6 on water, sanitation, and hygiene sets targets for universal access, where “universal” is explicitly stated to include healthcare facilities (United Nations Department of Economic and Social Affairs, 2023). Shortly after recognition under the Sustainable Development Goals, the World Health Organization sponsored a 2015 landscape report on environmental health services access in low- and middle-income countries (World Health Organization, 2015), which was the first major attempt at evidence synthesis and highlighted widespread coverage gaps.

Since the launch of this landscape report in 2015, there has been considerable mobilization of resources and political will for improving environmental health services. In 2018, the UN Secretary General issued a global call to action to ensure sustainable environmental health services in all healthcare facilities (United Nations Secretary-General, 2018). In 2019, the Joint Monitoring Program (JMP) of the World Health Organization (WHO) and United Nations Children’s Fund (UNICEF), which monitors progress towards Sustainable Development Goal 6 targets, released its first report on healthcare facilities (World Health Organization and United Nations Children’s Fund, 2019). Also in 2019, member states at the 72^nd^ World Health Assembly unanimously passed a resolution calling for action on environmental health services in healthcare facilities (World Health Assembly, 2019). In 2023, the United Nations General Assembly adopted resolution A/78/130, “Sustainable, safe and universal water, sanitation, hygiene, waste, and electricity services in health-care facilities,” which recognizes the importance of these services for the health and safety of patients and healthcare workers and aims to advance progress toward universal access (UN General Assembly, 2023). These actions have been accompanied by commitments from national governments and non-governmental development partners (see e.g., (World Health Organization and United Nations Children’s Fund (UNICEF), 2024) leading to a wide variety of national policies and guidelines, and mobilizing hundreds of millions of dollars for environmental health services in healthcare facilities.

Since 2015, programming and policy developments have often outpaced available evidence. For example, indicators for global monitoring basic service levels were not defined until 2019 (World Health Organization and United Nations Children’s Fund, 2019), and more nuanced indicators for monitoring advanced service levels remain unspecified. One of the centerpiece programmatic approaches developed by the WHO and UNICEF—the Water and Sanitation for Health Facility Improvement Tool (WASH FIT) (World Health Organization, 2022)—has become the most popular programmatic approach globally with implementation in over 70 countries (World Health Organization and United Nations Children’s Fund, 2025), and yet no conclusive evidence exists to document its effectiveness (Lineberger et al., 2025). WHO and UNICEF monitoring data indicate that only 36% of countries have costed implementation roadmaps at the national level that spell out a strategy for reaching universal coverage, in part due to lack of strong cost evidence and guidance on how to estimate costs at the national scale (World Health Organization and United Nations Children’s Fund, 2025).

Efficiently and sustainably advancing the global agenda on environmental health services in healthcare facilities will require a strong evidence base. Yet locating evidence for decision-making is difficult for several reasons. Because environmental health services as a cohesive target within global development are relatively recent, there are few attempts at evidence synthesis across the domain. Systematic reviews exist on a small number of niche topics, but have frequently failed to draw actionable conclusions, largely due to a lack of high-quality studies (Bouzid et al., 2018; Gozdzielewska et al., 2023; Watson et al., 2019).

Furthermore, research on environmental health services has historically been conducted across multiple disciplines that use different vocabularies and methods (e.g., medicine, public health, environmental engineering), which hinders attempts to locate and synthesize evidence for decision making (Anderson et al., 2025a).

### 1.1. Objectives

We conducted a literature inventory and created an interactive evidence map of our findings to consolidate and organize the breadth of available literature on environmental health services in healthcare facilities across disciplines. We aimed to capture all peer-reviewed literature on environmental health services, from cross-sectional studies on infrastructure access or staff knowledge, attitudes, and practices, to intervention trials, to operational and implementation science research. We restricted our focus to studies published in 2008 or later, the year the first major global guideline document defining the domain space of environmental health services was published by the WHO (Adams et al., 2008). Our research questions were as follows: (1) To what extent have environmental health services in healthcare facilities in LMICs been studied to date, and (2) how could this evidence inform policy and practice?

Our purpose was to curate available evidence into an interactive, consolidated, and organized repository for two primary reasons: one, so that policy makers and practitioners can begin to locate and apply the best available evidence for decision making, and two, so that researchers can see the broad landscape of available evidence and craft targeted research agendas that address the most pressing evidence gaps and avoid duplication of prior efforts. We chose a literature inventory approach (which extracts limited high-level information from a greater number and diversity of studies) rather than a systematic review (which extracts more detailed information from a narrower set of studies) to achieve this purpose.

We hypothesized that our evidence map would reveal an uneven distribution of evidence, leaving substantial gaps that will impede decision making. We specifically anticipated (1) a large proportion of the evidence to be baseline assessments documenting coverage of environmental health services but providing little other information, (2) an uneven distribution of evidence across environmental health services, with hygiene represented most often, and (2) predominance of cross-sectional study designs, even for intervention research where these designs are a substantial weakness.

This paper describes the methods and results of the literature inventory, describes the contents and usability of the interactive evidence map, and presents a case study of how the evidence map can be used to guide research and practice using sanitation as an example.

### 1.2. Scope and policy framework

We defined environmental health services—and therefore the scope of our search— based on the services monitored under the WHO/UNICEF Joint Monitoring Program and by the services included in the WHO’s 2008 environmental standards for healthcare facilities (Adams et al., 2008; Joint Monitoring Programme, 2024). Table 1 presents the environmental health services we included in our scope, along with their associated definitions.

**Table 1.**
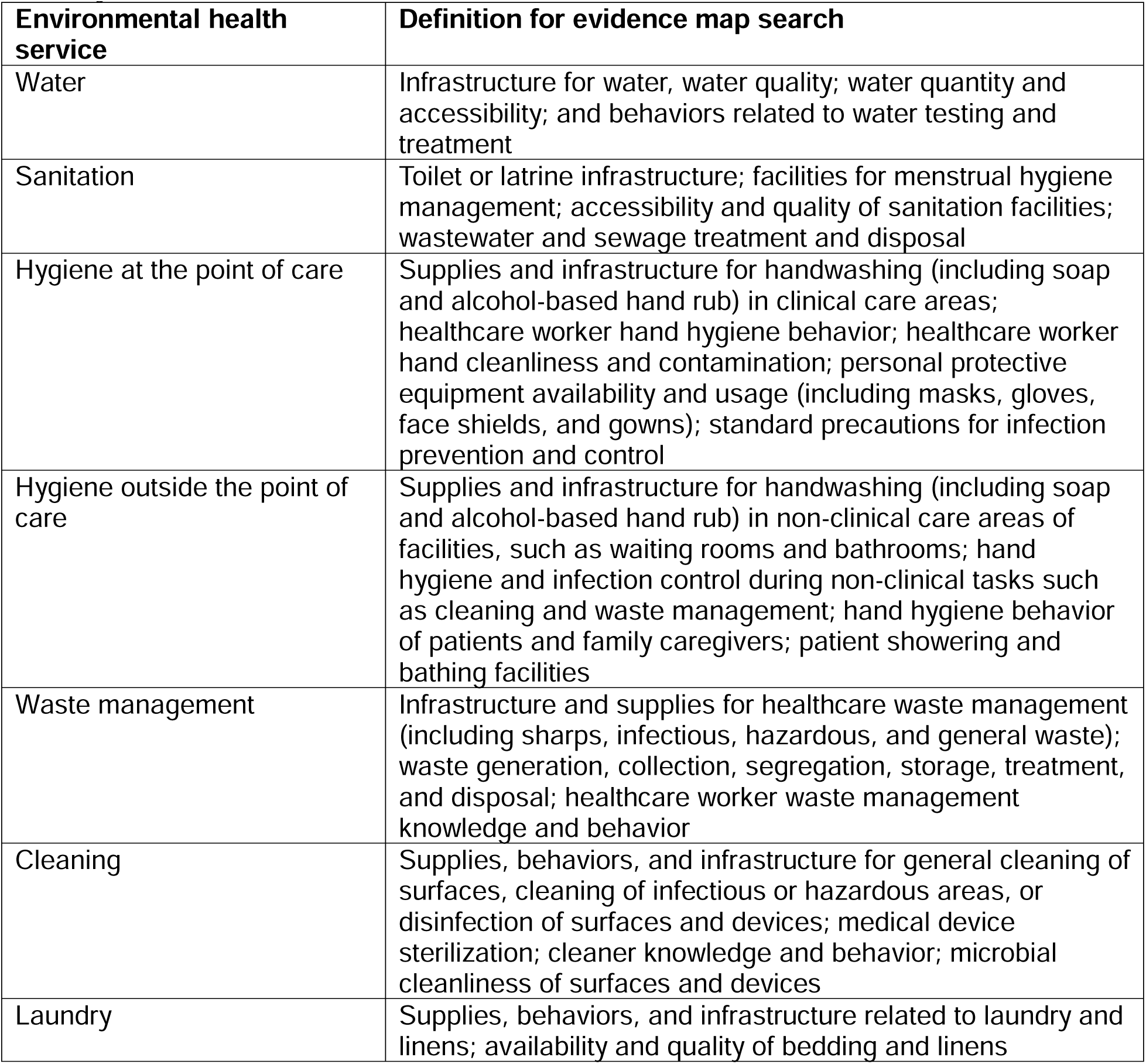
Definitions of environmental health services included in the evidence map and literature inventory.

| **Environmental health service** | **Definition for evidence map search** |
| --- | --- |
| Water | Infrastructure for water, water quality; water quantity and accessibility; and behaviors related to water testing and treatment |
| Sanitation | Toilet or latrine infrastructure; facilities for menstrual hygiene management; accessibility and quality of sanitation facilities; wastewater and sewage treatment and disposal |
| Hygiene at the point of care | Supplies and infrastructure for handwashing (including soap and alcohol-based hand rub) in clinical care areas; healthcare worker hand hygiene behavior; healthcare worker hand cleanliness and contamination; personal protective equipment availability and usage (including masks, gloves, face shields, and gowns); standard precautions for infection prevention and control |
| Hygiene outside the point of care | Supplies and infrastructure for handwashing (including soap and alcohol-based hand rub) in non-clinical care areas of facilities, such as waiting rooms and bathrooms; hand hygiene and infection control during non-clinical tasks such as cleaning and waste management; hand hygiene behavior of patients and family caregivers; patient showering and bathing facilities |
| Waste management | Infrastructure and supplies for healthcare waste management (including sharps, infectious, hazardous, and general waste); waste generation, collection, segregation, storage, treatment, and disposal; healthcare worker waste management knowledge and behavior |
| Cleaning | Supplies, behaviors, and infrastructure for general cleaning of surfaces, cleaning of infectious or hazardous areas, or disinfection of surfaces and devices; medical device sterilization; cleaner knowledge and behavior; microbial cleanliness of surfaces and devices |
| Laundry | Supplies, behaviors, and infrastructure related to laundry and linens; availability and quality of bedding and linens |

Our literature inventory and evidence map were guided by a policy framework developed by the WHO and UNICEF: *Water, Sanitation and Hygiene in Healthcare Facilities: Practical Steps to Achieve Universal Access to Quality Care* (hereafter called the “Eight Practical Steps”) (WHO/UNICEF, 2019). This document recommends eight action-oriented steps that national governments and other actors supporting environmental health services in healthcare facilities should follow to reach universal access. Table 2 defines the steps. The document was published in 2019 and has since heavily influenced WHO/UNICEF publications and country-level action.

**Table 2.** Summary of the Eight Practical Steps recommended by the WHO and UNICEF for country action to achieve universal access to environmental health services in healthcare facilities (WHO/UNICEF, 2019).

| **Step** | **Key actions** |
| --- | --- |
| Step 1: Conduct situation analysis and assessment | Assess governance structures, funding, and environmental and health policies (situation analysis)<br>Determine levels of environmental health services coverage and compliance with policies (assessment) |
| Step 2: Set targets and define roadmap | Define the national approach, targets, and intervention areas<br>Assign roles and responsibilities of stakeholders<br>Develop a costed roadmap for implementing the chosen approach |
| Step 3: Establish national standards and accountability mechanisms | Define contextualized national standards<br>Provide guidance on the design, costing, implementation, and operation of environmental health services<br>Create mechanisms to ensure that facilities meet established standards |
| Step 4: Improve and maintain infrastructure | Construct and/or rehabilitate infrastructure to meet standards<br>Develop policies and strategies for operations and maintenance<br>Ensure sufficient resources for operation and maintenance |
| Step 5: Monitor and review data | Establish systems for routine data collection and review<br>Analyze data to measure progress and hold stakeholders accountable |
| Step 6: Develop health workforce | Train and mentor the health workforce on environmental health services and infection prevention and control practices |
| Step 7: Engage communities | Engage communities in ongoing review of data on environmental health services coverage and implementation; engage communities in health and environmental health policies<br>Coordinate with community environmental health service providers where appropriate |
| Step 8: Conduct operational research and share learning | Document approaches, challenges, and solutions<br>Assess innovative approaches for potential for scale-up<br>Reflect on and revise programmatic strategies based on evidence and data |

For example, the 2024 WHO/UNICEF Global Framework for Action—which outlines the most pressing challenges for reaching universal access, proposes key next steps, and publishes commitments from governments and non-governmental actors—emphasizes the Eight Practical Steps as the underlying framework for action (World Health Organization and United Nations Children’s Fund (UNICEF), 2024).

Given its influence, we used the Eight Practical Steps framework as a core organizing principle for our literature inventory and evidence synthesis. We hypothesized that different types of information would be needed by policy makers and practitioners for each of the Eight Practical Steps. For instance, costing studies are needed for Step 2 on developing costed roadmaps; intervention trials are needed for Step 4 on implementing evidence-based interventions. For all studies included in our evidence map, we have cataloged it based on which of the Eight Practical Steps it might inform. We envision that this will help users better leverage the evidence map for practical decision making.

## 2. Methods

This evidence map is reported with reference to the “RepOrting Standards for Systematic Evidence Syntheses in environmental research” (ROSES) for systematic maps (Haddaway et al., 2018).

### 2.1. Search strategy

We originally searched PubMed (National Library of Medicine/National Institutes of Health), Scopus (Elsevier), and Global Health (EBSCOHost) databases on June 21, 2023, for studies that described environmental health services in healthcare facilities in LMICs. We conducted several pilot searches and refined search terms to yield an appropriate breadth of evidence. During these pilot searches, we assessed the comprehensiveness of the search by identifying several articles that would meet study inclusion criteria and verifying whether the search had captured these articles.

Our final search strategy included three clusters of terms: one cluster each for environmental health services, healthcare facilities, and LMICs. We applied Boolean logic such that search results had to contain at least one term from each cluster. Environmental health services encompassed the services listed in Table 1. Healthcare facility terms included general and specialized facility types (e.g., maternal healthcare settings). We drew LMIC country names from the 2022-2023 World Bank country income level definitions (World Bank, 2023). The search identified studies containing these terms at the title/abstract level. The search was restricted to articles published in English in 2008 or later, as the first WHO guidance for environmental health services in healthcare facilities was established in 2008 (Adams et al., 2008).

We conducted a search update on January 8, 2025. The search update also included a broadened set of LMIC-related search terms and adapted the Cochrane Effective Practice and Organisation of Care (EPOC)’s LMIC search filter (Cochrane Effective Practice and Organisation of Care (EPOC), 2021) to identify studies that took place in LMIC settings but did not specify a particular country. The search strategy for the search update is available in Supplementary Tables 1-3.

To supplement the database search, we conducted a hand search of three journals that commonly publish environmental health-related research but are not consistently indexed in databases: Journal of Water and Health; Journal of Water, Sanitation, and Hygiene for Development; and H2Open Journal. For hand-searching, we performed a search of each journal website using the keywords “Healthcare facility” or “Hospital.” We reviewed the title and abstract of each search result and exported the metadata of any references published in English between 2008 and the present, meeting other review inclusion criteria. Hand searches took place in February 2025.

### 2.2. Study selection

#### 2.2.1. Inclusion and exclusion criteria

We used the PECO (Population, Exposure, Comparator, Outcome) framework to identify eligible studies (Morgan et al., 2018). Studies were eligible if they described environmental health services in healthcare facilities in LMIC settings. Inclusion and exclusion criteria are detailed below.

For the population, we included any healthcare facility in an LMIC. We excluded studies that did not take place in a healthcare facility or that took place solely in a high-income country. We defined healthcare facilities as permanent institutions whose primary purpose is to provide direct patient care. Non-clinical settings within healthcare facilities (e.g., laboratories, sterilization processing units, waiting rooms), medical education settings, and assessments of healthcare worker behavior were eligible. Our definition of healthcare facilities excluded settings where medical care might be provided but was not the primary purpose, such as schools or group homes, and long-term rehabilitation facilities. We excluded facilities with the primary or sole purpose of providing dental care and nonpermanent facilities and field settings, such as temporary facilities established for vaccination campaigns, mobile clinics, ambulances, and community health workers. We defined LMICs using the 2022-2023 World Bank country income classifications (World Bank, 2023).

For the exposure, we included environmental health services as defined in Table 1. We included studies regardless of whether they focused on “hardware” (e.g., physical infrastructure, consumable products, and supplies), “software” (e.g., hygiene promotion, behavior change, training), or implementation strategies and operational research related to these services. We included studies focusing on sharps injuries only if they linked the injury to a problem with waste management (e.g., lack of safe disposal) or to personal protective equipment behavior (e.g., glove use), rather than to a clinical procedure. We excluded studies that did not examine any of the environmental health services defined in Table 1.

For comparison, we included any intervention or exposure comparator, as well as observational studies without a comparator. We had no exclusion criteria for comparison.

For outcomes, we included any outcome. We had no exclusion criteria by outcome. We included all study types that performed all primary research studies except case reports from a single patient. We included systematic and narrative reviews but excluded editorials and opinion pieces, clinical guidelines, or study protocols with no data collection or results. We also excluded mathematical modeling studies or laboratory-based simulation studies that did not collect or utilize data from a healthcare facility.

#### 2.2.2. Screening strategy

We used machine learning to prioritize studies for manual screening. For this process, senior members of the research team manually title-abstract screened an initial set of 1,000 randomly selected studies from our search. Of these, we identified 182 studies that met our inclusion criteria, while the remaining studies were excluded. These studies served as training data for machine learning using the Document Classification and Topic Extraction Resource (DoCTER) tool (ICF International, Inc., 2017). DoCTER uses machine learning algorithms to predict the relevance of unclassified citations based on text similarities in the titles and abstracts of a training dataset. Unclassified citations are stratified by predicted relevance, with probability scores for each. Previous studies have applied this machine learning approach in systematic reviews (Cawley et al., 2025; Rich et al., 2025).

All studies predicted to be relevant by machine learning were prioritized for manual screening. To mitigate the potential for missing relevant studies misclassified by machine learning, an additional 1,000 studies with the highest probability score among those not predicted as relevant were also manually screened. The remaining studies that were predicted not to be relevant were excluded without being manually screened. Each citation was independently screened at the title-abstract level by two reviewers. If their decisions conflicted, a third senior research team member reviewed the title and abstract to resolve the conflicts. We restricted reviewers from screening studies that they had authored. We used Covidence software (Veritas Health Innovation, n.d.) to manage references during the screening process.

### 2.3. Metadata extraction and tagging

For studies included at the title-abstract level, we exported metadata (unique ID number, abstract, and citation information) from Covidence into an Excel spreadsheet. To categorize studies for the evidence map, we performed a literature inventory tagging process that involved assigning tags based on information contained in the title and abstract. Tags fell into four categories: environmental health service topic, study design, policy/practice relevance, and special topics. For policy/practice relevance, we mapped each study on to at least of the Eight Practical Steps, based on the type of information collected (e.g., costing studies were tagged with Step 2 on developing costed roadmaps; intervention studies were tagged with Step 4 on improving environmental health services).

Each study received at least one tag from the environmental health services, study design, and policy/practice relevance categories; special topics tags were applied only if applicable. Studies could receive multiple tags from each category (e.g., if they examined multiple environmental domains or used a mixed-methods approach). If studies were missing data related to one or more tags, they could be categorized under an “other/not specified” option. The research team developed and refined tag categories and definitions through a process of discussion and iteration. We piloted the tagging process on an initial set of 50 studies to refine definitions and improve reviewer consistency. Supplementary Table 4 provides the complete list of tags and their corresponding definitions.

For studies identified in the original search, research team members worked in groups of two or three to separately tag studies, compare tags, and resolve discrepancies through discussion. During the search update, team members independently tagged studies. In both cases, a senior team member performed quality control by reviewing tagged studies, verifying that tags had been applied accurately, and making necessary adjustments. Initially, we performed this quality control process for all studies. When a team member reached at least 90% accuracy in their tagging, we shifted to a spot-check approach, performing quality control for one in five tagged studies. Throughout the tagging process, we continually refined tag definitions as new study types emerged.

In addition to tags applied by researchers, we classified the geographic locations where studies were conducted using an automated tagging procedure. We used a Python script to detect names of LMICs in study titles and/or abstracts and subsequently performed manual data cleaning to correct or add country names if missing.

### 2.4. Study quality assessment

We did not conduct a study quality assessment. Our aim was to characterize the landscape of evidence on environmental health services in healthcare facilities. As such, the evidence map encompasses a broad scope of literature and does not delve into the findings or quality of individual studies. We collected and reported information on study design through the study tagging process; however, study design alone may not accurately reflect the strength or quality.

### 2.5. Data synthesis and presentation

We compiled the included studies, along with their tags, in a database. Using study tags as the unit of analysis, we generated descriptive statistics for the number and proportion of studies associated with each tag and with combinations of tags. We created graphical depictions of the study database using a Tableau data visualization dashboard. Graphics included heat maps of the number of studies categorized within each environmental domain, policy/practice relevance category, and study design category. The dashboard also includes graphics showing the number of studies published annually from 2008 to 2025, the journals in which they were published, and the geographic locations where they were conducted. Using the dashboard, we performed a narrative synthesis of the trends in environmental health services research. The data visualization dashboard is <u>accessible on Tableau Public</u>.

## 3. Results

### 3.1. Search results

The initial search and search update yielded 62,814 citations, of which 25,436 were duplicates. After excluding articles via machine learning (n = 24,011), we identified 13,367 for manual screening. Hand-searching yielded 36 additional articles. During the tagging process, we identified 848 articles that had been retained in the screening stage but did not meet eligibility criteria, resulting in a total of 4,381 studies included in the evidence map. The ROSES flow diagram (Figure 1) summarizes search and screening results.

**Figure 1.**
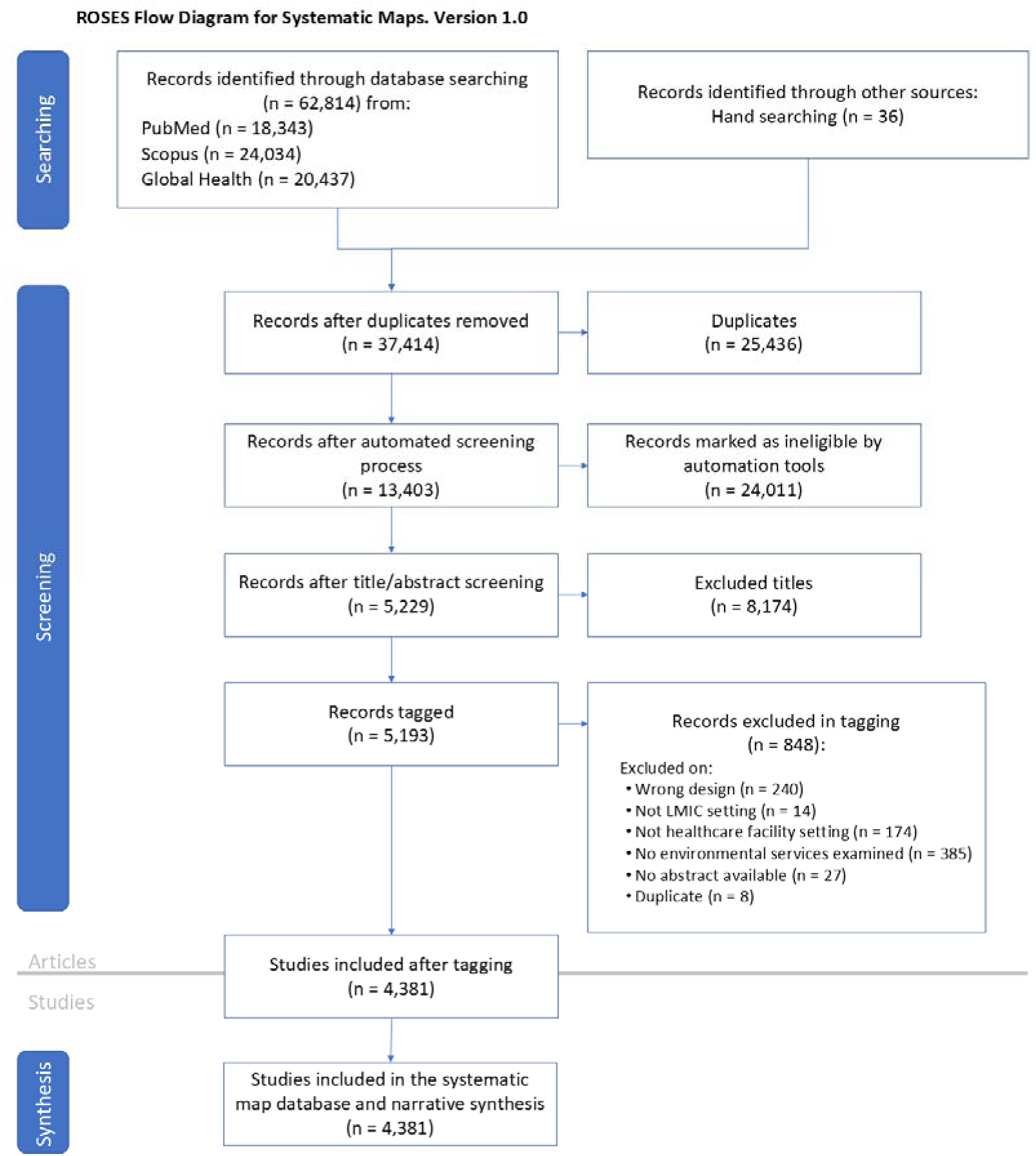
ROSES flow diagram for studies included in a systematic evidence map of environmental health services in healthcare facilities in low- and middle-income countries, 2025.

### 3.2. Evidence map user interface

The Tableau dashboard consolidates information on environmental health services into an interactive interface (Figure 2). Users can select a heatmap cell, chart component, or map country to filter the dashboard by this element. In addition to viewing the filtered dashboard, users can view and download a list of the articles that are relevant to the selected element of interest – such as articles related to an environmental health service domain, study design, geographic area, or keyword search.

**Figure 2.**
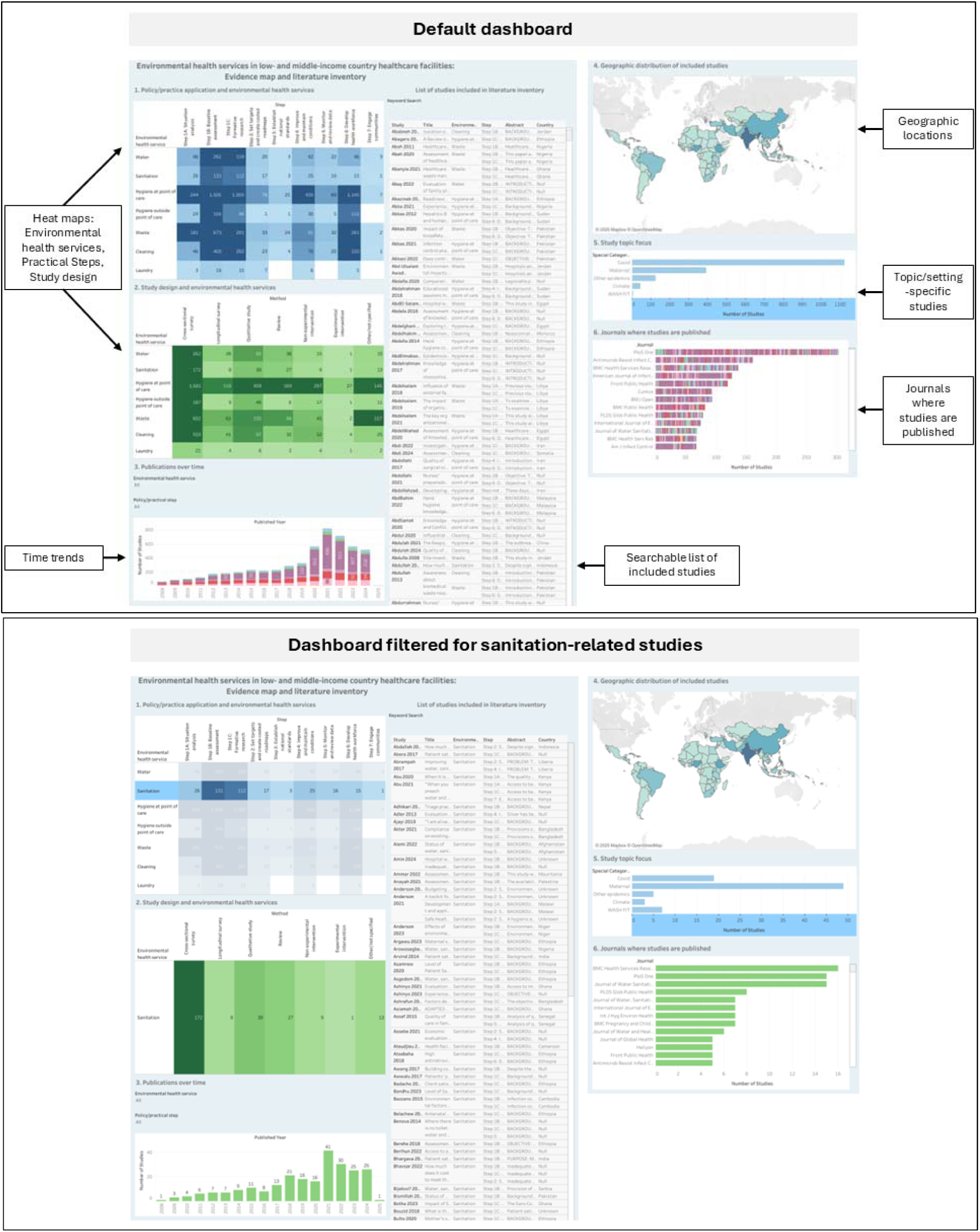
Tableau interactive dashboard for evidence map and literature inventory, including default view (top) and view when filtered to show studies related to sanitation (bottom).

Box 1 describes a hypothetical case study of how the evidence map may be used by practitioners and researchers.

#### Box 1.

Using the evidence map dashboard for research and practice: a hypothetical case study on sanitation.

##### Part 1. Context and objectives

This case study imagines that policymakers in a middle-income country are interested in developing a country-level sanitation improvement plan for healthcare facilities. They hope to create a plan that is appropriate for the local context, feasible to execute with available resources, and aligned with best practices for safe sanitation. The policymakers have partnered with a research team to assist in forming an evidence-based plan and developing an accompanying research agenda. The policy and research team is interested in gathering evidence to support their sanitation improvement planning process, including information on: (1) prior interventions that have been effective in improving sanitation in healthcare facilities; (2) considerations for program implementation, including likely barriers and facilitators to program delivery, workforce development needs, and monitoring and evaluation frameworks; (3) budget estimates; and (4) gaps and trends in published research on sanitation in healthcare facilities, to inform research agenda development.

##### Part 2. Application of the evidence map for policy and research objectives

The team uses the Tableau dashboard’s heat map of policy-practice applications and environmental health services to aid in their sanitation improvement planning process. They filter the heat map to review formative research on sanitation (Step 1C), providing information on factors that have been found to support or impede sanitation improvement to develop a contextualized theory of change. Then, they filter the heat map to review studies related to improving and maintaining sanitation conditions (Step 4) to identify prior sanitation interventions and understand how they were implemented. They filter by Step 2 and Step 6 to gather information about budgets and workforce development, respectively. The team uses the geographic map to prioritize studies from their country and neighboring countries with similar resource constraints, to maximize contextual relevance. They identify projects that have already been implemented and learn more about the actors involved in sanitation-related programs and research in their country. Finally, maternal and child health is a priority issue for several policymakers, and they are interested in highlighting this in the national improvement plan. The team filters the study topic chart to identify research on sanitation in maternal settings, providing insight into setting-specific considerations.

To develop a research agenda for sanitation, the team examines study designs used to assess sanitation in healthcare facilities. There are many cross-sectional surveys on sanitation (n=272). These may provide useful insight into existing conditions and indicate that further baseline assessment is not necessary. However, there are few sanitation interventions: the evidence map includes nine non-experimental intervention studies and one experimental intervention study focused on sanitation. Developing and evaluating interventions to improve sanitation in healthcare facilities – especially using experimental approaches – would be a valuable research contribution. Researchers also examine the trend in sanitation-related publications over time, finding a steady increase from 2008 through 2024. They can perform an in-depth review of the most recent studies to gain a clearer picture of the current state of the field. Later, after collecting and analyzing data, researchers use the dashboard to view the journals where sanitation-related studies are most often published to identify a suitable platform for their work.

### 3.3. Study design

Most studies (63%, n = 2,777) were cross-sectional surveys, involving the collection of quantitative data at a single point in time. Other observational studies employed longitudinal surveys with quantitative measurements at two or more time points (5%, n = 230) and/or qualitative methods (14%, n = 627). Five percent of studies (n = 205) employed a mixed-methods observational approach, incorporating both qualitative and quantitative data. Seven percent of the studies (n = 288) were reviews, including narrative reviews, systematic reviews, and meta-analyses.

Among 397 studies that measured changes related to an intervention or quality improvement program, 92% (n = 364) employed a non-experimental or quasi-experimental design, such as a before-and-after, difference-in-differences, or time-series design. Eight percent of intervention evaluation studies (n = 33) employed an experimental design involving a randomized controlled trial. Most randomized controlled trials (82%, n = 27) were related to hygiene at the point of care. Four randomized controlled trials (12%) pertained to cleaning, and one to two pertained to each of the other environmental health services (Figure 3).

**Figure 3.**
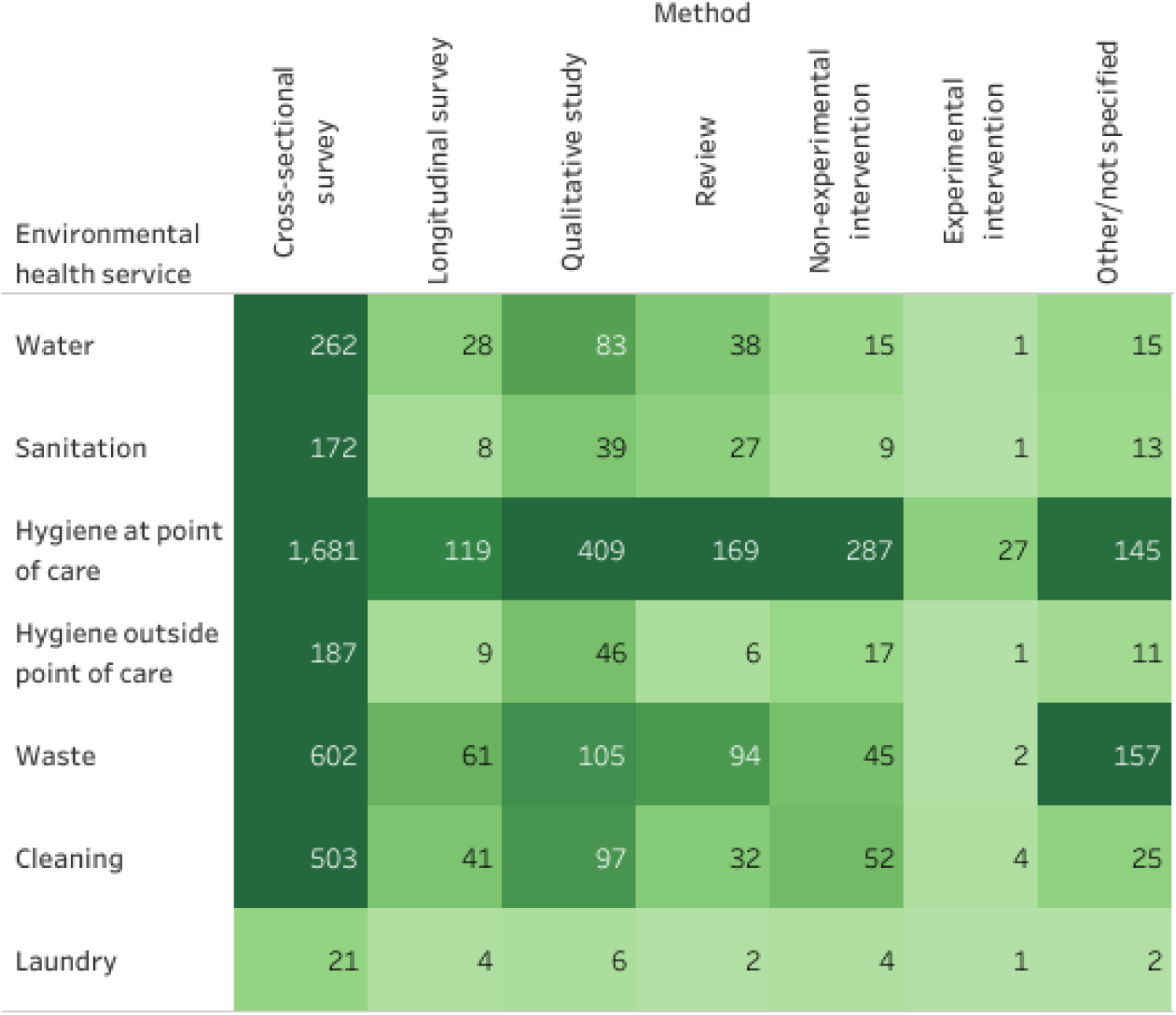
Heat map of methods and environmental health service topics for 4,381 studies on environmental health services in LMIC healthcare facilities. Studies that involved multiple methods and/or topics are included in all applicable heat map cells.

### 3.4. Environmental health service topics

There was substantial variation in the amount of research related to each environmental health service. The most commonly studied service was hygiene at the point of care, which was nearly three times as prevalent as the next most common service, waste management. Water, hygiene outside of points of care, sanitation, and laundry were each tagged in fewer than 10% of studies (Table 3).

**Table 3.**
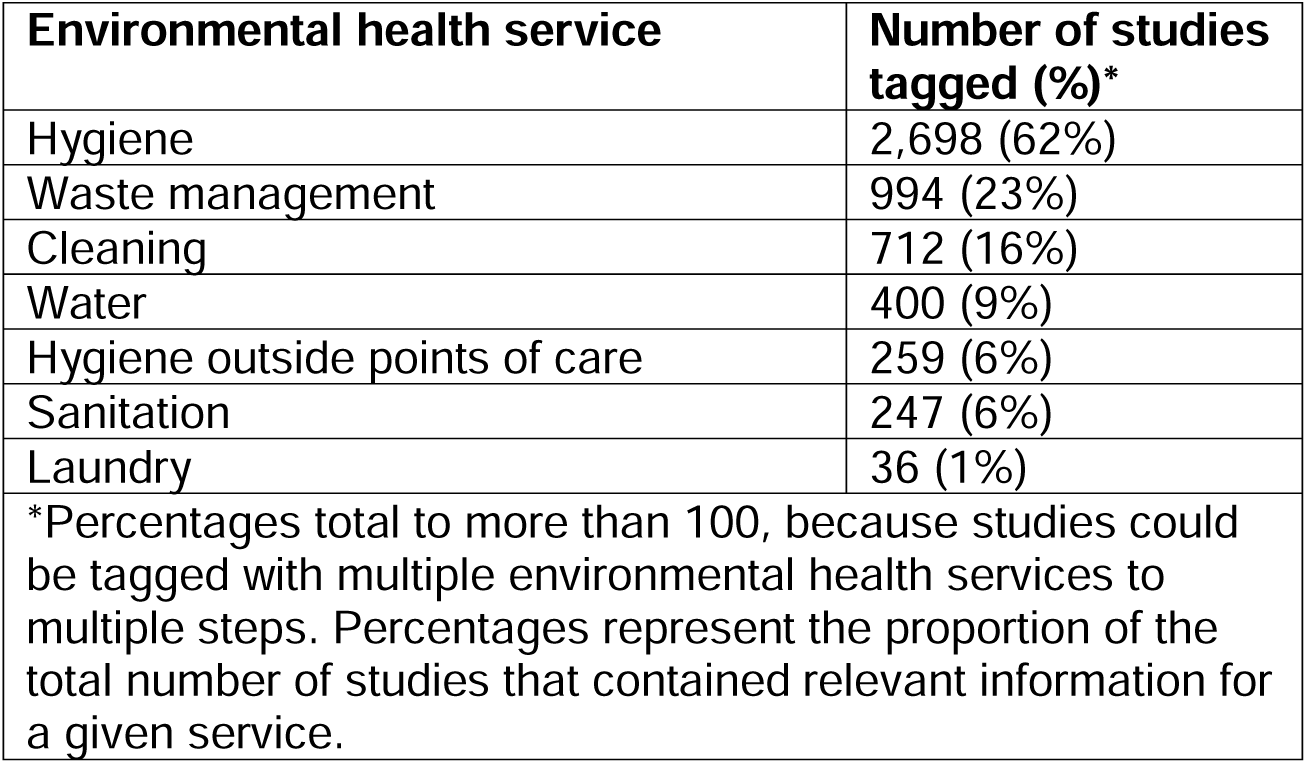
Distribution of environmental health services reported within the evidence map.

Most studies were published in scientific journals in the public health, infection prevention and control, or WASH sectors. PLOS One, a general scientific journal, was the most common journal overall, publishing 157 evidence map studies. Among hygiene- and cleaning-focused studies, the most prevalent journals were *PLOS One*, *BMC Health Services Research*, *American Journal of Infection Control*, *Antimicrobial Resistance and Infection Control* and *International Journal of Environmental Research and Public Health*. Water- and sanitation-related studies were also commonly published in *Journal of Water, Sanitation, and Hygiene for Development*. Among waste management studies, the most prevalent journals were *Waste Management & Research* and *Waste Management*.

### 3.5. Policy and practice relevance: Practical Steps framework

We examined the relationship of studies to policy and guideline development, program evaluation, and practice using a framework derived from the WHO/UNICEF Eight Practical Steps (World Health Organization, 2019) (Figure 4).

**Figure 4.**
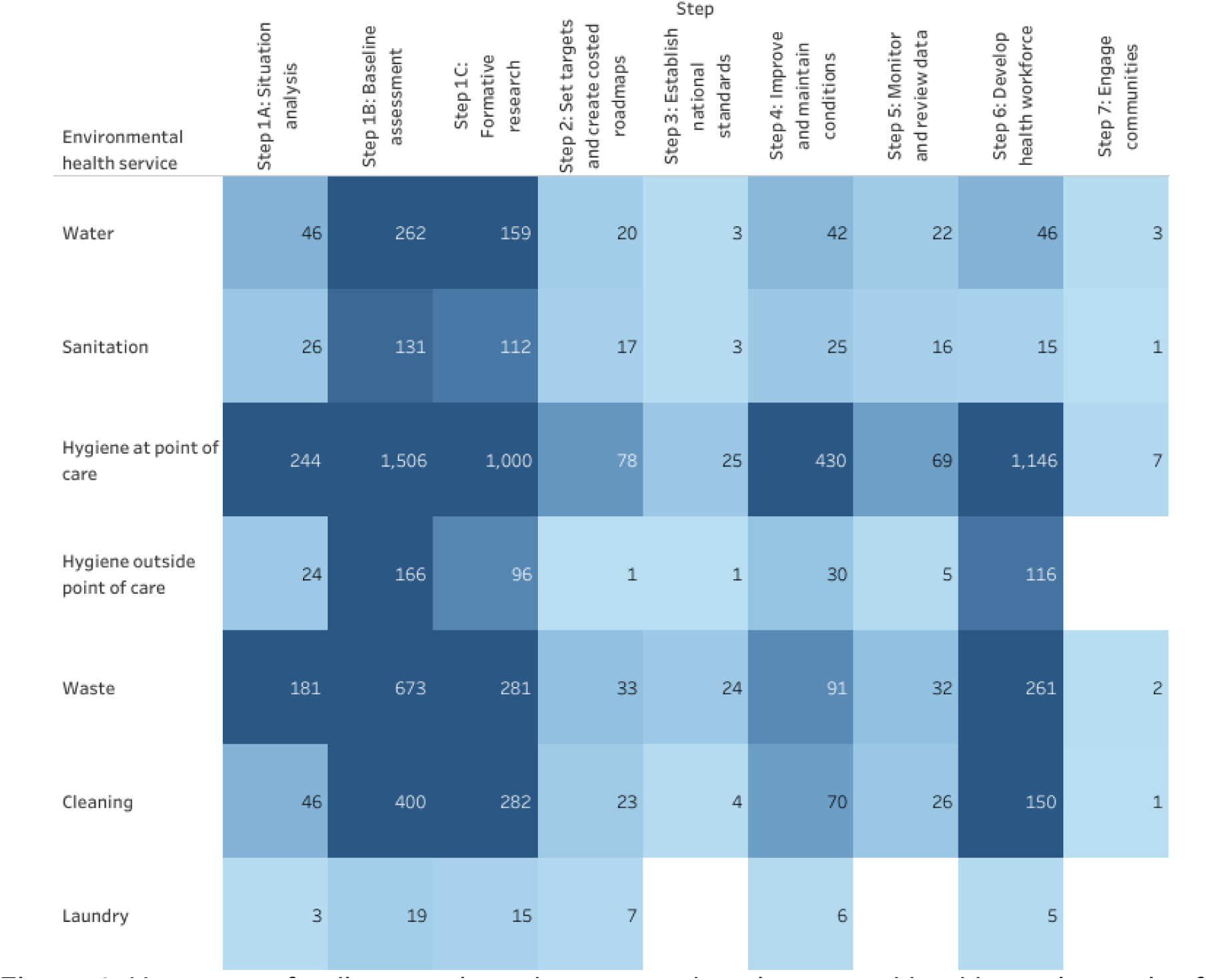
Heat map of policy-practice relevance and environmental health service topics for 4,381 studies on environmental health services in LMIC healthcare facilities. Studies that involved multiple practical steps and/or topics are included in all applicable heat map cells.

Fifty-two percent of studies contained information relevant to multiple steps. More than half of studies (58%) contained baseline assessments (Step 1B). Approximately a third of studies were tagged for Step 1C (formative research that either described barriers and determinants of safe environmental health services, or that linked environmental health services to outcomes such as behavior change, care delivery, or health) and Step 6 (health workforce development). Other Steps were rarely represented, with each being tagged for fewer than 15% of studies (Table 4).

**Table 4.** Prevalence of tags for the Eight Practical Steps within the evidence map.

| Practical step | Number of studies tagged (%)* |
| --- | --- |
| Step 1A: Situation analysis | 474 (11%) |
| Step 1B: Baseline assessments | 2,548 (58%) |
| Step 1C: Formative research | 1,596 (36%) |
| Step 2: Set targets and coordination | 118 (3%) |
| Step 3: Establish national standards | 52 (1%) |
| Step 4: Improve and maintain conditions | 574 (13%) |
| Step 5: Monitor and review data | 121 (3%) |
| Step 6: Develop health workforce | 1,484 (34%) |
| Step 7: Engage communities | 10 (0.2%) |
| *Percentages total to more than 100, because studies could be assigned to multiple steps. Percentages represent the proportion of the total number of studies that contained relevant information for a given step. |  |

*Step 1A – Conduct situation analysis:* Among studies examined aspects of the policy context or enabling environment for environmental health services in healthcare facilities, 51% (n=244) examined hygiene outside the point of care, and 38% (n=181) examined waste management. Forty-five percent of situation analyses were cross-sectional assessments, and 18% (n=86) were reviews.

*Step 1B – Conduct baseline assessment* Most baseline assessments (80%, n = 2,049) involved the collection of cross-sectional survey data. Of these studies, 10% (n = 252) were also categorized under Step 1A, as they examined both the baseline conditions and the broader enabling environment of the healthcare facility. Most baseline assessments focused on hygiene at the point of care (59%, n=1506), followed by waste (26%, n=673) and cleaning (16%, n=400). Less than 10 percent of baseline assessments described water, sanitation, laundry, or hygiene outside the point of care. Seven percent of baseline assessments (n=179) took place in maternal, newborn, or child healthcare settings, and 24% (n=620) described conditions in the context of the COVID-19 pandemic.

*Step 1C – Formative research:* Among studies examining barriers, facilitators, or downstream impacts of environmental health services, 528 (33%) included a baseline assessment, and 123 (8%) included a situation analysis. Most formative research studies employed quantitative methods to examine the associations between environmental health services and other exposures or outcomes, with 72% (n = 1148) using a cross-sectional or longitudinal survey approach. Twenty-three percent of formative research studies (n = 365) employed qualitative methods to gather stakeholder perspectives on the barriers to and impacts of implementing safe environmental health services.

*Step 2 – Set targets and establish a coordination mechanism:* Two-thirds (n=78) of the studies describing target-setting, costing and budgeting, and coordination pertained to hygiene at the point of care. Fewer studies described these processes in relation to waste (28%, n=33), cleaning (19%, n=23), water (17%, n=20), sanitation (14%, n=17), laundry (6%, n=7), or hygiene outside the point of care (1%, n=1). We identified 20 studies that described targets, budgeting, or coordination in the context of intervention implementation or operations and maintenance.

*Step 3 – Establish national WASH or healthcare waste management standards:* Most studies describing the development or implementation of national standards were focused on hygiene at the point of care (48%, n=25) or waste management (46%, n=24). Approximately half of these studies were also tagged under Step 1A, as they assessed the enabling environment for services in addition to describing national-level standards.

*Step 4 – Improve and maintain environmental health services:* Three-quarters of studies evaluating interventions, quality improvement programs, or implementation strategies pertained to hygiene at the point of care (n=430), followed by waste management (16%, n=91) and cleaning (12%, n=70). Fifty-nine percent of studies tagged under Step 4 (n=338) used a quasi-experimental evaluation approach to evaluate changes related to an intervention, while 9% (n=53) used cross-sectional surveys and 8% (n=44) used qualitative methods.

*Step 5 – Monitor and review data:* Among 121 studies that utilized routine monitoring systems to evaluate environmental health services, 36% (n=43) were cross-sectional surveys and 48% (n=58) were longitudinal surveys. Most of these studies (57%, n=69) evaluated hygiene at the point of care.

*Step 6 – Develop a health workforce:* Of the studies tagged under Step 6, 67% (n = 347) were also categorized as baseline assessments. Twenty-three percent (n=347) of Step 6 studies were also categorized under Step 4 (“improve and maintain environmental health services”). Across all study designs in the Step 6 category, 77% of studies (n = 1,146) examined hygiene behaviors at the point of care. Eighteen percent of studies (n=261) described knowledge or practices related to waste management, 10% (n=150) related to cleaning, and less than 10% each related to water, sanitation, laundry, or hygiene outside the point of care.

*Step 7 – Engage communities:* Very few studies (n = 10, 0.2%) employed community-based participatory methods. Of the studies that employed these methods, 50% took place in a maternal, newborn, or child health setting, and 70% used a qualitative data collection approach.

### 3.6 Geographic distribution

Studies took place in 109 LMICs. Studies most frequently originated from India (n = 624, 14%), Ethiopia (n = 335, 8%), Nigeria (n = 334, 8%), China (n = 285, 6%), and Iran (n = 260, 6%). Of the 132 countries classified as LMICs by the World Bank, 55 (42%) had at least 10 studies identified in the evidence map, while 26 (19%) had no identified studies. Most of the LMICs with no identified studies were small island nations in the Caribbean or Pacific (e.g., Dominica, St. Lucia, Marshall Islands, Samoa) or countries in Europe and Central Asia (e.g., Azerbaijan, Belarus, Moldova, Montenegro). Figure 5 depicts the distribution of research across countries.

**Figure 5.**
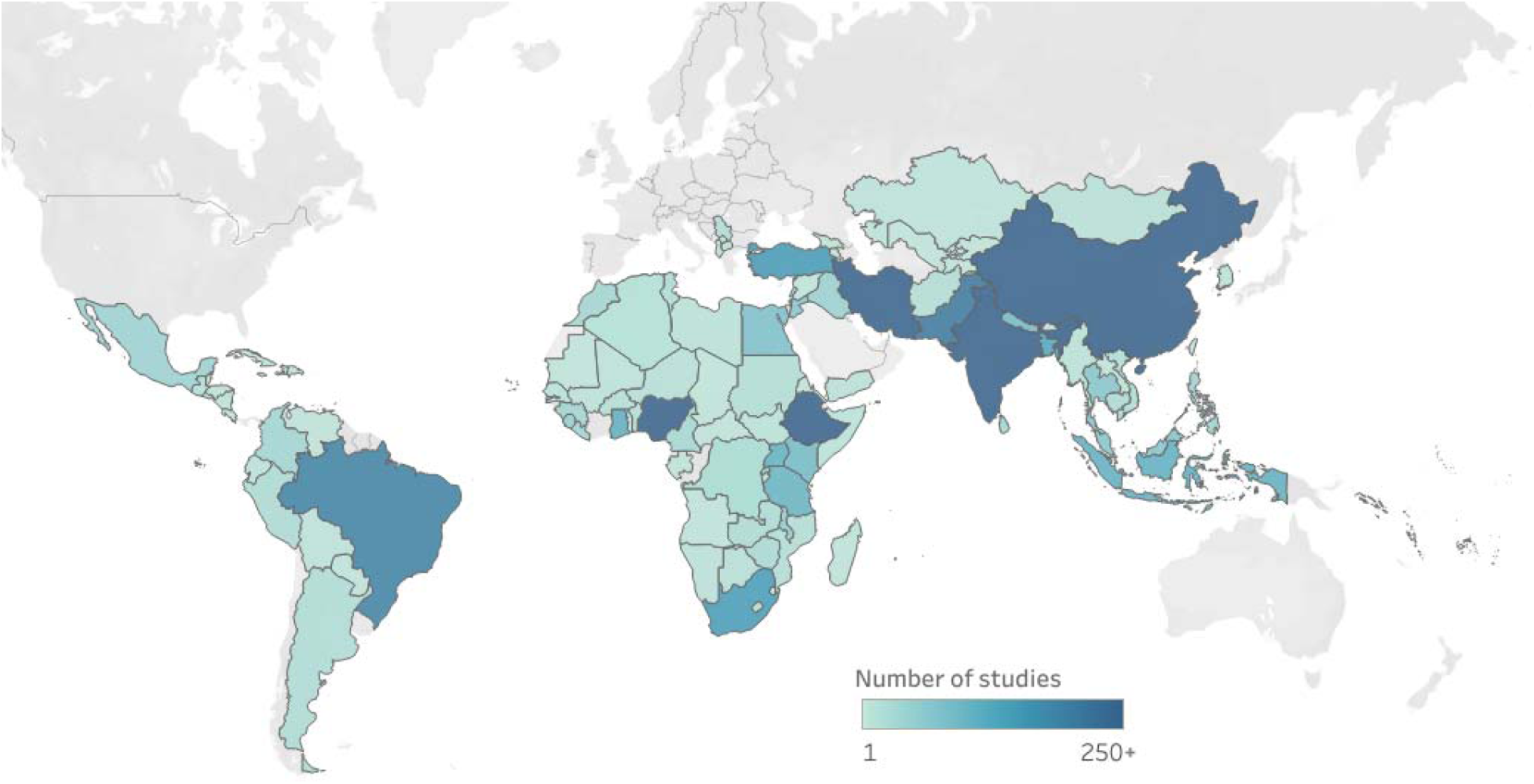
Geographic distribution of 4,381 studies on environmental health services in healthcare facilities across low- and middle-income countries. A darker color indicates a larger number of identified studies.

### 3.7. Publication time trends

We examined the number of studies published annually between 2008 and 2025 and found an overall upward trend in publications during this period. Fewer than 100 studies were published annually between 2008 and 2011, compared with 410 in 2024. The number of publications increased sharply starting in 2020, reaching a peak in 2021. This upward trend was driven in part by studies examining hygiene at the point of care during the COVID-19 pandemic. Forty-six percent of studies published in 2020, and 54% of those published in 2021, described environmental health services during the COVID-19 pandemic or examined repercussions of COVID-19 for services. In 2024, the proportion of studies pertaining to COVID-19 decreased to 29%. As the search update took place in January 2025, data for the number of studies published in 2025 are incomplete. Figure 6 illustrates the annual number of studies published overall and for each environmental service domain.

**Figure 6.**
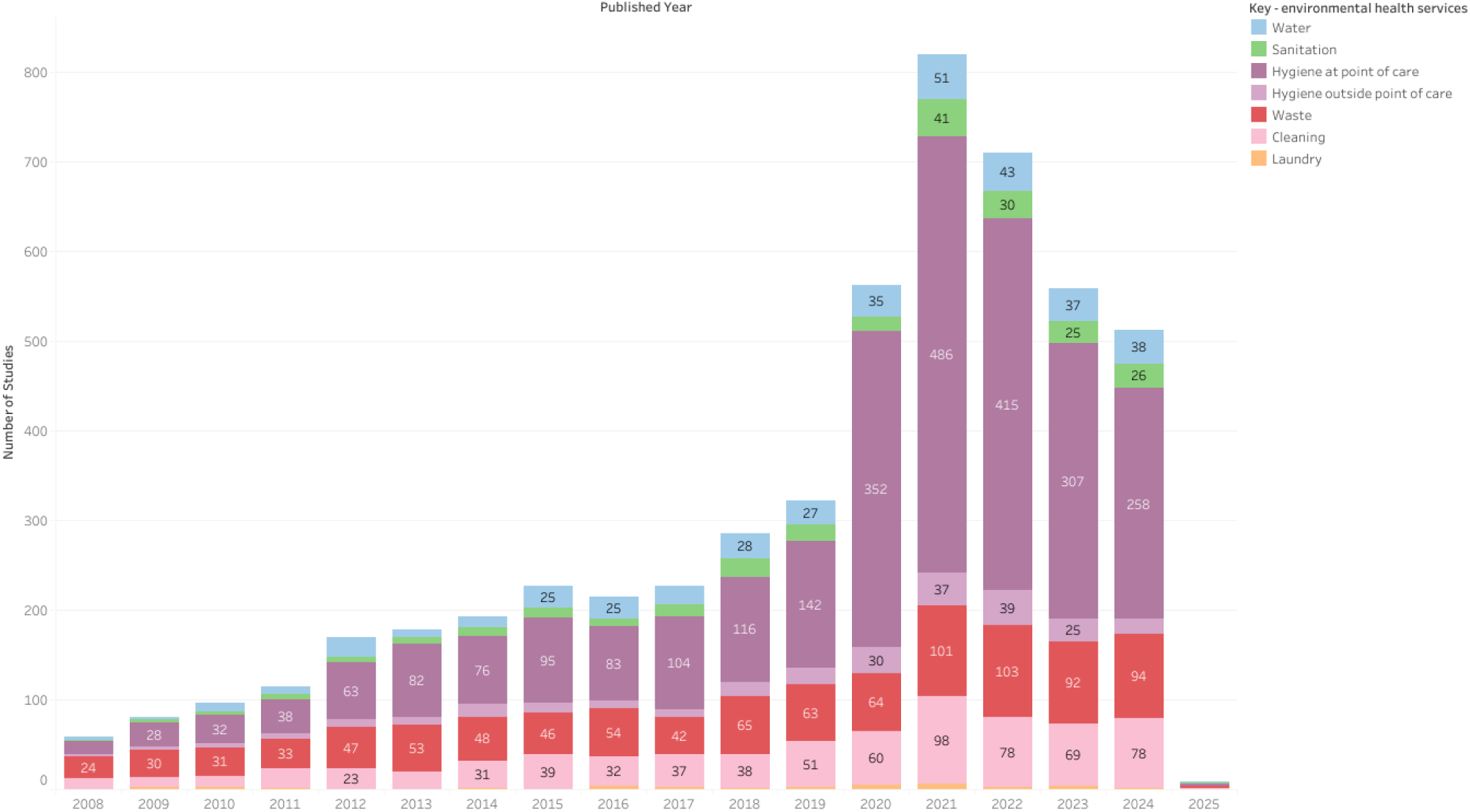
Time trend in the annual number of studies published on environmental health services in LMIC healthcare facilities.

### 3.8. Priority topics and populations

Nine percent of studies (n = 392) were conducted primarily in maternal, newborn, and child health settings or examined environmental health-related outcomes among maternal, newborn, or under-five populations. Many of these studies examined environmental health services in maternity wards or measured the role of environmental health services in maternal patient satisfaction and quality of care. Twenty-seven percent of studies (n = 1,169) contained themes related to the COVID-19, including studies that took place in specialized COVID-19 care wards or that described COVID-19 risks and precautions in general healthcare settings. Nearly all these studies (88%) pertained to hygiene at the point of care, describing hand hygiene and infection control services and behaviors related to the COVID-19 pandemic. The majority (64%) of COVID-19-related studies were cross-sectional surveys.

Three percent of studies (n=141) took place in the context of other disease epidemics and outbreaks, such as Ebola, Lassa fever, or influenza. Forty-seven studies (1%) pertained to aspects of climate change or climate resilience, including studies that examined healthcare facilities’ preparedness for extreme weather events, described effects of extreme weather on infrastructure, or reported on environmental sustainability of services.

We identified 10 studies that reported on the use of the WASH FIT quality improvement tool in healthcare facilities (World Health Organization, 2022). These studies applied WASH FIT to describe the status of environmental health services, plan and implement improvements, and assess progress toward targets. Two WASH FIT-related studies were also conducted in COVID-19 treatment facilities, while others were conducted in general healthcare settings.

## 4. Discussion

We compiled a literature inventory and evidence map that synthesized over 4,000 articles on environmental health services in LMIC healthcare facilities published since 2008. Our goal was to catalog the broad spectrum of peer-reviewed evidence—including all study designs and outcomes—to create a database applicable to research, policy, and practice. We organized this evidence according to the Eight Practical Steps framework from the WHO and UNICEF, based on how study information could support decision-making for each step.

The evidence base on environmental health services in healthcare facilities has accelerated since 2008, with an increasing number of new studies published each year. This reflects a growing recognition of environmental health services as a dedicated domain space within global health and development. This is also likely influenced by the COVID-19 pandemic, as we observed a noticeable spike beginning in 2020 that corresponded with an increase in our COVID-related keyword tags. However, while publication numbers declined from 2022 to 2024, they remain considerably above 2019 levels, suggesting ongoing increases in investment independent of COVID-19.

While the overall quantity of evidence has increased substantially, its distribution across topics remains uneven. Our evidence map highlights areas that are well saturated with evidence to support evidence-based decision-making and those where sparse evidence is likely to hinder progress. We undertook this literature inventory and evidence map in part because we hypothesized that the current evidence base on environmental health services is unevenly distributed across topics. We specifically anticipated (1) a large proportion of the evidence would consist of baseline assessments documenting coverage of environmental health services but providing little other information, (2) evidence would be unevenly distributed across environmental health services, with hygiene represented most often, and (3) cross-sectional designs would be most prevalent, even for intervention research where these designs are a substantial weakness.

We found our hypothesis to be largely correct. We hypothesized that a large proportion of studies would be baseline assessments, and that evidence supporting other steps within the Eight Practical Steps framework would be sparse. Our results support this. Forty-one percent of all studies were solely baseline assessments (Step 1B) that reported coverage levels of environmental services but did not conduct a situation assessment of the broader context or link services to inputs or outcomes. A majority (71%) of these baseline assessment studies were published after 2015. Studies pre-2015 and shortly thereafter were valuable in informing the Sustainable Development Goals and setting the stage for subsequent global action. However, we argue that dedicating continued effort and resources to these types of baseline assessments a decade later will yield diminishing returns. These studies often simply conclude what is already known—that environmental health services in LMICs lag substantially behind universal access targets and require action to improve them. An estimated 53% of countries had completed baseline assessments in 2015 as part of national monitoring systems that can be used for prioritizing targeted areas for service improvement under Step 1 (World Health Organization and United Nations Children’s Fund, 2025). Future resources would be better spent strengthening these systems rather than conducting ad hoc, independent studies using the same indicators.

We hypothesized that evidence would be unevenly distributed across different environmental health services. Our results also support this. A majority (62%) of the included articles focused on hand hygiene at clinical points of care, and fewer than 10% focused on water, sanitation, hygiene outside of points of care, or laundry. This likely reflects the differing disciplinary histories of research on environmental health services. Hand hygiene at points of care is a key component of infection prevention and control efforts for decades, and by extension has been the subject of medical research during that time. Conversely, global health and development efforts around water and sanitation have historically been led by non-medical disciplines that have not engaged with healthcare facility settings (Anderson et al., 2025a) This is reflected in the disciplinary breakdown of journals for evidence from each of the environmental health services. Many of the identified studies on hand hygiene and cleaning were published in journals focused on infection control or clinical practices, whereas studies on water and sanitation were more commonly published in environmental health- or WASH-related journals.

Finally, we hypothesized that cross-sectional study designs would predominate, and our evidence map corroborated this. Most (63%) studies used cross-sectional quantitative surveys for data collection. In total, we identified only 33 experimental intervention studies, of which 82% were for point-of-care hygiene interventions. Reliance on cross-sectional data undermines the evidence base, particularly for Steps 4 and 6 on improving and maintaining conditions and developing the health workforce. Evidence-based decision making for these steps will rely on studies that robustly evaluate intervention effectiveness, and cross-sectional studies are ill-suited to this type of evaluation. Steps 4 and 6 likely represent the largest area of investment across the Eight Practical Steps, in part due to the costs of skilled personnel and infrastructure construction, operations, and maintenance; yet the evidence base supporting these steps is relatively weak compared to other steps.

### 4.1. Limitations

While we conducted a broad literature search across several databases, supplemented by hand-searching, it may have missed relevant studies published in non-indexed journals or those whose titles and abstracts did not contain search keywords. Notably, our search did not examine grey literature. We expect that evidence supporting Steps 2, 3, and possibly 5 is more likely to appear in the grey literature than in other steps, in part because they cannot be as easily studied using classic public health study designs. We did not search for grey literature primarily because we were unable to identify a feasible search strategy. A key challenge was that environmental health services are distributed across a wide variety of disciplines, and there are few databases that support complex searches and bulk export of results with standardized bibliographic information, which was essential given the breadth of our search and screening. Difficulty of searching grey literature reflects a need for better options to index applied, policy-relevant evidence so that it can be used by researchers and practitioners.

The search and screening strategy was also restricted to English-language studies, potentially excluding some research from countries where publication in languages other than English is more common. We expect this may impact our results on the geographic coverage of studies. However, we expect our other key findings (e.g., the high proportion of baseline assessments and hygiene studies) would be affected, as these factors are uncorrelated with the language in which studies were published.

We did not perform full-text extraction of the articles included, as our objective was to compile and describe a comprehensive literature inventory. This study, therefore, does not provide insight into study findings, quality, or intervention effect sizes. Further systematic reviews could focus on specific dimensions of environmental health services in healthcare facilities, such as a single service domain, sub-population, study type, or geographic region, for in-depth data extraction and analysis.

### 4.2. Next steps for strengthening global evidence on environmental health services in healthcare facilities

#### 4.2.1. Pivot from baseline assessments to intervention, operational, and policy research

The need for environmental health services is well established, and we expect that new baseline assessments will add little value, especially where they measure the presence/absence of services following JMP indicators. Research into developing new indicators for advanced service levels would be beneficial, as the JMP currently lacks such indicators. While baseline assessments under Step 1 are well established, situation analyses— which explore the nuanced social, political, economic, geographic, and other contextual factors that enable and inhibit service delivery—are uncommon. This may be in part because well-established tools exist for baseline assessments but not situation analyses (Anderson et al., 2025b).

Beyond Step 1, we found a need to strengthen evidence to support all remaining steps, either because of a lack of evidence and/or weak study designs. Steps 4 and 6 (improve conditions and strengthen health workforce) can be largely evaluated by classical study designs familiar to public health and medical researchers (e.g., randomized controlled trials, quasi-experimental studies). Other steps will require engaging disciplines that have historically been less engaged in environmental health sciences research, such as economics, political science, and public policy. However, methods do not necessarily need to be complicated. Qualitative studies that simply document how a particular policy process occurred, the actors involved, and the success drivers (see, e.g., an example documenting the process of developed costed roadmaps in Nepal (Chettry et al., 2024)) can provide valuable evidence where little currently exists.

#### 4.2.2. Research water, sanitation, and other under-studied environmental health services

While hand hygiene and waste management still undoubtedly warrant further study, we found an order of magnitude less research on water, sanitation, hygiene outside points of care, and laundry. Water in particular is fundamental to cleaning, hand hygiene, and direct provision of medical care, making the literature gap especially important. Some research on water and sanitation from household, school and other non-healthcare settings may be translatable—for example studies on how climate and hydrogeologic conditions affect infrastructure operations and maintenance. However, other aspects such as management models for water and sanitation systems that involve a unique set of health and non-health system actors will require specific study.

#### 4.2.3. Use stronger study designs for interventions research

Intervention studies are important but costly evidence. We argue that targeted investment in experimental designs for some environmental health services would be beneficial (e.g., on interventions to improve operations, maintenance, and sustainability of water, sanitation, and waste management infrastructure). However, quasi-experimental designs will also be important and less costly. Assessments could leverage secondary data from health information systems or national surveys to assess changes in key output, outcome, and impact measures over time associated with rollout of national or subnational programs. Some countries track water, sanitation, and hygiene service levels in healthcare facilities in health information systems, which can be used for evaluation purposes, though data availability and quality vary (Chatterley et al., 2018). Healthcare associated infections are challenging to track but incorporated into some national health information management systems. Other indicators like workforce retention, patient fees and revenue, and patient satisfaction are other candidate indicators that may be tracked through information systems or other national monitoring surveys that would be suitable for evaluations (Bergh et al., 2022; Lopez et al., 2020; Wagenaar et al., 2016). When available, these databases can serve as a resource for understanding current service levels and tracking change over time (Dubik et al., 2024). Government health authorities should consider incorporating environmental health service indicators into health information systems to facilitate the routine monitoring of service delivery.

#### 4.2.4. Conduct targeted systematic reviews and build research agendas by topic

One of the key purposes of our literature inventory and evidence map was to organize the landscape of research on environmental health services to guide future research and maximize its utility for decision making. Our findings suggest some high-level guidance (e.g., saturation of baseline assessments, relatively high proportion of hygiene at point of care research), but further exploration will be needed. For topics that have a high concentration of studies, systematic review on specific outcomes can extract further information on effectiveness, contextual barriers and facilitators, and other key information that were beyond the scope of our literature inventory but is essential for evidence-based decision making. For topics lacking studies, research agendas can be developed to guide primary research and fill identified gaps.

## 5. Conclusion

This systematic evidence map reveals that, although a substantial body of evidence exists on environmental health services in healthcare facilities, coverage is uneven across environmental health services, study types, and relevance to steps within the Eight Practical Steps policy framework. Hygiene and infection control have been extensively studied, and environmental health services have been characterized in baseline assessments. However, there are comparatively few formative research studies and intervention studies related to other environmental health services, and even less evidence on policy and financing. A lack of scientific evidence poses a barrier to improving services and monitoring progress toward global targets. In addition to conducting further primary research on under-studied areas, evidence synthesis can help the field identify common knowledge gaps and research priorities. This study has compiled a literature inventory that can serve as a basis for additional, targeted systematic reviews. Reviews could focus on specific environmental domains (e.g., water or sanitation), geographic regions (e.g., fragile contexts), or clinical settings (e.g., maternal healthcare) to evaluate research trends in these areas and inform future directions. Systematic reviews can serve as a basis for developing research agendas; input from non-research collaborators can ensure that these agendas are relevant to programmatic priorities and practical constraints (Yoshida et al., 2016). Coordination among research, policy, and program stakeholders can support the translation of research findings into global progress toward universal access to safe environmental health services.

## Data Availability

All data produced in the present work are contained in the manuscript.

## Acknowledgements

We thank the UNC undergraduate researchers who contributed to title-abstract screening and/or literature inventory tagging: Abby Arcuri, Ablah Siddiq, Aiden Keller, Alexandra Ayala, Anna Pederson, Arjun Suresh, Avi Rathod, Caroline Jones, Chauncey Chen, Chidinma Okoye, Chris Zou, Claire Macdaid, Jessica Li, Joseph Sullivan, Kailey Wadsworth, Layan Qamari, Lucy Fang, Mang Iang, Manir Hede, Mercy Adekola, Michael Fehl, Rachitha Vijayakumar, Sari Ghirmay-Morgan, Scout Correa, Shreeya Patel, Simone Rothaput, Vijay Seethana, Vivian Campbell, and Zhilin Zhai. We also thank Sophie Guo, Samiira Hassan, and Sophia Shen for their assistance in developing the Tableau data visualization dashboard. We thank Michelle Cawley for providing advice on literature search and screening strategies.

