## Supplementary Information for "Mapping the evidence on environmental health services in healthcare facilities in low- and middle-income countries: A systematic literature inventory of over 4,000 studies"

**Contents**


1. Search strategy for database search update (Supplementary tables 1-3)
2. Tag categories for evidence map of environmental health services in healthcare facilities (Supplementary table 4)


**Search strategy for database search update run January 8, 2025**

***Supplementary table 1. PubMed (National Library of Medicine / National Institutes of Health): January 8, 2025***

| Search | Query | Number of Results |
|---|---|---|
| 1 | ("standard precautions"[tiab] OR "infection control"[tiab] OR "infection prevention"[tiab] OR IPC[tiab] OR "personal protective equipment"[tiab] OR PPE[tiab] OR "splash protection"[tiab] OR "respiratory protection"[tiab] OR mask[tiab] OR masks[tiab] OR glove[tiab] OR gloves[tiab] OR gown[tiab] OR gowns[tiab] OR goggles[tiab] OR "eye protection"[tiab] OR "face shield"[tiab] OR "face shields"[tiab] OR respirator[tiab] OR respirators[tiab] OR water[tiab] OR sanitation[tiab] OR sanitary[tiab] OR plumbing[tiab] OR sewage[tiab] OR sewer*[tiab] OR latrine[tiab] OR latrines[tiab] OR toilet[tiab] OR toilets[tiab] OR hygiene[tiab] OR hygienic[tiab] OR shower[tiab] OR showers[tiab] OR soap[tiab] OR soaps[tiab] OR detergent[tiab] OR detergents[tiab] OR handwashing[tiab] OR "hand washing"[tiab] OR "hand hygiene"[tiab] OR laundry[tiab] OR bedsheet[tiab] OR bedsheets[tiab] OR bedding[tiab] OR linen[tiab] OR linens[tiab] OR waste[tiab] OR wastes[tiab] OR landfill[tiab] OR landfills[tiab] OR dump[tiab] OR dumps[tiab] OR drainage[tiab] OR wastewater[tiab] OR "waste water"[tiab] OR disposal[tiab] OR disinfect*[tiab] OR cleaner[tiab] OR cleaners[tiab] OR cleaning[tiab] OR cleanliness[tiab] OR janitor*[tiab] OR housekeep*[tiab]) | 1,688,229 |
| 2 | (healthcare[tiab] OR "health care"[tiab] OR hospital[tiab] OR hospitals[tiab] OR clinic[tiab] OR clinics[tiab] OR "health facility"[tiab] OR "health facilities"[tiab] OR "health center"[tiab] OR "health centers"[tiab] OR healthcenter[tiab] OR healthcenters[tiab] OR healthcentre[tiab] OR healthcentre[tiab] OR "health centre"[tiab] OR "health centres"[tiab] OR "health post"[tiab] OR "health post"[tiab] OR healthpost[tiab] OR healthposts[tiab] OR "health setting"[tiab] OR "health settings"[tiab] OR "medical facility"[tiab] OR "medical facilities"[tiab] OR "medical center"[tiab] OR "medical centers"[tiab] OR "medical centre"[tiab] OR "medical centres"[tiab] OR "medical post"[tiab] OR "medical posts"[tiab] OR "medical setting"[tiab] OR "medical settings"[tiab] OR "delivery facility"[tiab] OR "delivery facilities"[tiab] OR "delivery center"[tiab] OR "delivery centers"[tiab] OR "delivery centre"[tiab] OR "delivery | 2,689,559 |

| Search | Query | Number of Results |
| --- | --- | --- |
| | centres"[tiab] OR "delivery clinic"[tiab] OR "delivery clinics"[tiab] OR "birth facility"[tiab] OR "birth facilities"[tiab] OR "birth center"[tiab] OR "birth centers"[tiab] OR "birth centre"[tiab] OR "birth centres"[tiab] OR "birth clinic"[tiab] OR "birth clinics"[tiab] OR "maternal facility"[tiab] OR "maternity facility"[tiab] OR "maternal facilities"[tiab] OR "maternity facilities"[tiab] OR "maternal center"[tiab] OR "maternity center"[tiab] OR "maternal centers"[tiab] OR "maternity centers"[tiab] OR "maternal centre"[tiab] OR "maternal centers"[tiab] OR "maternity clinic"[tiab] OR "maternity clinics"[tiab] OR "maternal clinic"[tiab] OR "maternal clinics"[tiab] OR "maternity home"[tiab] OR "maternity homes"[tiab] OR "maternity waiting homes"[tiab] OR "dispensary"[tiab] OR "dispensaries"[tiab]) | |
| 3 | Afghanistan[tiab] OR Algeria[tiab] OR Angola[tiab] OR Anguilla[tiab] OR Antigua[tiab] OR Barbuda[tiab] OR Argentina[tiab] OR Armenia[tiab] OR Armenian[tiab] OR Aruba[tiab] OR Azerbaijan[tiab] OR Bahamas[tiab] OR Bahrain[tiab] OR Bangladesh[tiab] OR Barbados[tiab] OR Benin[tiab] OR Byelarus[tiab] OR Byelorussian[tiab] OR Belarus[tiab] OR Belorussian[tiab] OR Belorussia[tiab] OR Belize[tiab] OR Bhutan[tiab] OR Bolivia[tiab] OR Botswana[tiab] OR Brazil[tiab] OR Brunei[tiab] OR "Burkina Faso"[tiab] OR "Burkina Fasso"[tiab] OR "Upper Volta"[tiab] OR Burundi[tiab] OR Urundi[tiab] OR Cambodia[tiab] OR "Khmer Republic"[tiab] OR Kampuchea[tiab] OR Cameroon[tiab] OR Cameroons[tiab] OR Cameron[tiab] OR Camerons[tiab] OR "Cape Verde"[tiab] OR "Cayman Islands"[tiab] OR "Central African Republic"[tiab] OR Chad[tiab] OR Chile[tiab] OR China[tiab] OR Colombia[tiab] OR Comoros[tiab] OR "Comoro Islands"[tiab] OR Comores[tiab] OR Mayotte[tiab] OR Congo[tiab] OR Zaire[tiab] OR "Cook Islands"[tiab] OR "Costa Rica"[tiab] OR "Cote d'Ivoire"[tiab] OR "Ivory Coast"[tiab] OR Croatia[tiab] OR Cuba[tiab] OR Cyprus[tiab] OR Djibouti[tiab] OR "French Somaliland"[tiab] OR Dominica[tiab] OR "Dominican Republic"[tiab] OR "East Timor"[tiab] OR "East Timur"[tiab] OR "Timor Leste"[tiab] OR Ecuador[tiab] OR Egypt[tiab] OR "United Arab Republic"[tiab] OR "El Salvador"[tiab] OR Eritrea[tiab] OR Ethiopia[tiab] OR "Falkland Islands"[tiab] OR "Las Malvinas"[tiab] OR Fiji[tiab] OR Gabon[tiab] OR "Gabonese Republic"[tiab] OR Gambia[tiab] OR Gaza[tiab] OR "Georgia Republic"[tiab] OR "Georgian Republic"[tiab] OR Ghana[tiab] OR "Gold Coast"[tiab] OR Greece[tiab] OR Grenada[tiab] OR Guatemala[tiab] OR Guinea[tiab] OR Guam[tiab] OR Guadeloupe[tiab] OR Guiana[tiab] OR Guyana[tiab] OR Haiti[tiab] OR Honduras[tiab] OR "Hong Kong"[tiab] OR India[tiab] OR Maldives[tiab] OR Indonesia[tiab] OR Iran[tiab] OR Iraq[tiab] OR Jamaica[tiab] OR Jordan[tiab] OR Kazakhstan[tiab] OR Kazakh[tiab] OR Kenya[tiab] OR Kiribati[tiab] OR Korea[tiab] OR Kosovo[tiab] OR Kuwait[tiab] OR Kyrgyzstan[tiab] OR Kirghizia[tiab] OR "Kyrgyz Republic"[tiab] OR Kirghiz[tiab] OR Kirgizstan[tiab] OR "Lao PDR"[tiab] OR Laos[tiab] OR Lebanon[tiab] OR Lesotho[tiab] OR Basutoland[tiab] OR Liberia[tiab] OR Libya[tiab] OR | 1,819,547 |

| Search | Query | Number of Results |
| --- | --- | --- |
| | Macau[tiab] OR Madagascar[tiab] OR "Malagasy Republic"[tiab] OR Maldives[tiab] OR Malaysia[tiab] OR Malaya[tiab] OR Malay[tiab] OR Sabah[tiab] OR Sarawak[tiab] OR Malawi[tiab] OR Nyasaland[tiab] OR Mali[tiab] OR Malta[tiab] OR "Marshall Islands"[tiab] OR Martinique[tiab] OR Mauritania[tiab] OR Mauritius[tiab] OR "Agalega Islands"[tiab] OR Mexico[tiab] OR Micronesia[tiab] OR "Middle East"[tiab] OR Mongolia[tiab] OR Montserrat[tiab] OR Morocco[tiab] OR Ifni[tiab] OR Mozambique[tiab] OR Myanmar[tiab] OR Myanma[tiab] OR Burma[tiab] OR Namibia[tiab] OR Nauru[tiab] OR Nepal[tiab] OR Niui[tiab] OR "Netherlands Antilles"[tiab] OR "New Caledonia"[tiab] OR Nicaragua[tiab] OR Niger[tiab] OR Nigeria[tiab] OR "Northern Mariana Islands"[tiab] OR Oman[tiab] OR Mayotte[tiab] OR Muscat[tiab] OR Pakistan[tiab] OR Palau[tiab] OR Palestine[tiab] OR Panama[tiab] OR Paraguay[tiab] OR Peru[tiab] OR Philippines[tiab] OR Philipines[tiab] OR Phillipines[tiab] OR Phillippines[tiab] OR Polynesia[tiab] OR "Puerto Rico"[tiab] OR Qatar[tiab] OR Reunion[tiab] OR Rwanda[tiab] OR Ruanda[tiab] OR "Saint Kitts"[tiab] OR "St Kitts"[tiab] OR Nevis[tiab] OR "Saint Lucia"[tiab] OR "St Lucia"[tiab] OR "Saint Vincent"[tiab] OR "St Vincent"[tiab] OR Grenadines[tiab] OR Samoa[tiab] OR "Samoan Islands"[tiab] OR "Navigator Island"[tiab] OR "Navigator Islands"[tiab] OR "Sao Tome"[tiab] OR "Saudi Arabia"[tiab] OR Senegal[tiab] OR Serbia[tiab] OR Montenegro[tiab] OR Seychelles[tiab] OR "Sierra Leone"[tiab] OR Singapore[tiab] OR "Sri Lanka"[tiab] OR Ceylon[tiab] OR "Solomon Islands"[tiab] OR Somalia[tiab] OR "South Africa"[tiab] OR Sudan[tiab] OR Suriname[tiab] OR Surinam[tiab] OR Swaziland[tiab] OR Syria[tiab] OR Tajikistan[tiab] OR Tadzhikistan[tiab] OR Tadjikistan[tiab] OR Tadzhik[tiab] OR Tanzania[tiab] OR Thailand[tiab] OR Togo[tiab] OR "Togolese Republic"[tiab] OR Tokelau[tiab] OR Tonga[tiab] OR Trinidad[tiab] OR Tobago[tiab] OR Tunisia[tiab] OR Turkey[tiab] OR Turkmenistan[tiab] OR Turkmen[tiab] OR "Turks Caicos"[tiab] OR "Turks and Caicos"[tiab] OR Tuvalu[tiab] OR Uganda[tiab] OR "United Arab Emirates"[tiab] OR Uruguay[tiab] OR Uzbekistan[tiab] OR Uzbek[tiab] OR Vanuatu[tiab] OR "New Hebrides"[tiab] OR Venezuela[tiab] OR Vietnam[tiab] OR "Viet Nam"[tiab] OR "Virgin Islands"[tiab] OR "West Bank"[tiab] OR Yemen[tiab] OR Yugoslavia[tiab] OR Zambia[tiab] OR Zimbabwe[tiab] OR “developing population*”[tiab] OR “developing world”[tiab] OR “less developed countr*”[tiab] OR “less developed nation*”[tiab] OR “less developed world”[tiab] OR “lesser developed countr*”[tiab] OR “lesser developed nation*”[tiab] OR “lesser developed world”[tiab] OR “under developed countr*”[tiab] OR “under developed nation*”[tiab] OR “under developed world”[tiab] OR “underdeveloped countr*”[tiab] OR “underdeveloped nation*”[tiab] OR “underdeveloped world”[tiab] OR “middle income countr*”[tiab] OR “middle income nation*”[tiab] OR “middle income population*”[tiab] OR “low income countr*”[tiab] OR “low income nation*”[tiab] OR “low income population*”[tiab] OR “lower income countr*”[tiab] OR “lower income | |

| Search | Query | Number of Results |
|---|---|---|
| | nation*"[tiab] OR "lower income population*"[tiab] OR "underserved countr*"[tiab] OR "underserved nation*"[tiab] OR "underserved population*"[tiab] OR "under served population*"[tiab] OR "under served nation*"[tiab] OR "under served population*"[tiab] OR "deprived countr*"[tiab] OR "deprived population*"[tiab] OR "high burden countr*"[tiab] OR "high burden nation*"[tiab] OR "countdown countr*"[tiab] OR "countdown nation*"[tiab] OR "poor countr*"[tiab] OR "poor nation*"[tiab] OR "poor population*"[tiab] OR "poor world"[tiab] OR "poorer countr*"[tiab] OR "poorer nation*"[tiab] OR "poorer population*"[tiab] OR "poorer world"[tiab] OR "developing econom*"[tiab] OR "less developed econom*"[tiab] OR "underdeveloped econom*"[tiab] OR "under developed econom*"[tiab] OR "middle income econom*"[tiab] OR "low income econom*"[tiab] OR "lower income econom*"[tiab] OR "low gdp"[tiab] OR "low gnp"[tiab] OR "low gross domestic"[tiab] OR "low gross national"[tiab] OR "lower gdp"[tiab] OR "lower gnp"[tiab] OR "lower gross domestic"[tiab] OR "lower gross national"[tiab] OR lmic[tiab] OR lmics[tiab] OR "third world"[tiab] OR "lami countr*"[tiab] OR "transitional countr*"[tiab] OR "emerging econom*"[tiab] OR "emerging nation*"[tiab] | |
| 4 | #1 AND #2 AND #3 | 23,064 |
| 5 | #4 AND (2008:2030[pdat]) | 19,012 |
| 6 | #5 AND (English[Language]) | 17,210 |
| **Total** | | **18,343** |

***Supplementary table 2. Scopus (Elsevier): January 8, 2025***

| Search | Query | Number of Results |
|---|---|---|
| 1 | TITLE-ABS ( "standard precaution" OR "standard precautions" OR "infection control" OR "infection prevention" OR IPC OR "personal protective equipment" OR PPE OR "splash protection" OR "respiratory protection" OR mask OR masks OR glove OR gloves OR gown OR gowns OR goggles OR "eye protection" OR "face shield" OR "face shields" OR respirator OR respirators OR water OR sanitation OR sanitary OR plumbing OR sewage OR sewer* OR latrine OR latrines OR toilet OR toilets OR hygiene OR hygienic OR shower OR showers OR soap OR soaps OR detergent OR detergents OR handwashing OR "hand washing" OR "hand hygiene" OR laundry OR bedsheet OR bedsheets OR bedding OR linen OR linens OR waste OR wastes OR landfill OR landfills OR dump OR dumps OR drainage OR wastewater OR "waste water" OR disposal OR disinfect* OR cleaners OR cleaning OR cleanliness OR janitor* OR housekeep* ) | 6,532,831 |
| 2 | TITLE-ABS ( healthcare OR "health care" OR hospital OR hospitals OR clinic OR clinics OR "health facilit*" OR "health center*" OR healthcenter* OR healthcentre* OR "health centre*" OR "health post*" | 3,492,583 |

| Search | Query | Number of Results |
|---|---|---|
| | OR healthpost* OR "health setting*" OR "medical facilit*" OR "medical center*" OR "medical centre*" OR "medical post*" OR "medical setting*" OR "delivery facilit*" OR "delivery center*" OR "delivery centre*" OR "delivery clinic*" OR "birth facilit*" OR "birth center*" OR "birth centre*" OR "birth clinic*" OR "matern* facilit*" OR "matern* center*" OR "matern* centre*" OR "matern* clinic*" OR dispensary OR dispensaries ) | |
| 3 | TITLE-ABS (Afghanistan OR Algeria OR Angola OR Anguilla OR Antigua OR Barbuda OR Argentina OR Armenia OR Armenian OR Aruba OR Azerbaijan OR Bahamas OR Bahrain OR Bangladesh OR Barbados OR Benin OR Byelarus OR Byelorussian OR Belarus OR Belorussian OR Belorussia OR Belize OR Bhutan OR Bolivia OR Botswana OR Brazil OR Brunei OR "Burkina Faso" OR "Burkina Fasso" OR "Upper Volta" OR Burundi OR Urundi OR Cambodia OR "Khmer Republic" OR Kampuchea OR Cameroon OR Cameroons OR Cameron OR Camerons OR "Cape Verde" OR "Cayman Islands" OR "Central African Republic" OR Chad OR Chile OR China OR Colombia OR Comoros OR "Comoro Islands" OR Comores OR Mayotte OR Congo OR Zaire OR "Cook Islands" OR "Costa Rica" OR "Cote d'Ivoire" OR "Ivory Coast" OR Croatia OR Cuba OR Cyprus OR Djibouti OR "French Somaliland" OR Dominica OR "Dominican Republic" OR "East Timor" OR "East Timur" OR "Timor Leste" OR Ecuador OR Egypt OR "United Arab Republic" OR "El Salvador" OR Eritrea OR Ethiopia OR "Falkland Islands" OR "Las Malvinas" OR Fiji OR Gabon OR "Gabonese Republic" OR Gambia OR Gaza OR "Georgia Republic" OR "Georgian Republic" OR Ghana OR "Gold Coast" OR Greece OR Grenada OR Guatemala OR Guinea OR Guam OR Guadeloupe OR Guiana OR Guyana OR Haiti OR Honduras OR "Hong Kong" OR India OR Maldives OR Indonesia OR Iran OR Iraq OR Jamaica OR Jordan OR Kazakhstan OR Kazakh OR Kenya OR Kiribati OR Korea OR Kosovo OR Kuwait OR Kyrgyzstan OR Kirghizia OR "Kyrgyz Republic" OR Kirghiz OR Kirgizstan OR "Lao PDR" OR Laos OR Lebanon OR Lesotho OR Basutoland OR Liberia OR Libya OR Macau OR Madagascar OR "Malagasy Republic" OR Maldives OR Malaysia OR Malaya OR Malay OR Sabah OR Sarawak OR Malawi OR Nyasaland OR Mali OR Malta OR "Marshall Islands" OR Martinique OR Mauritania OR Mauritius OR "Agalega Islands" OR Mexico OR Micronesia OR "Middle East" OR Mongolia OR Montserrat OR Morocco OR Ifni OR Mozambique OR Myanmar OR Myanma OR Burma OR Namibia OR Nauru OR Nepal OR Niui OR "Netherlands Antilles" OR "New Caledonia" OR Nicaragua OR Niger OR Nigeria OR "Northern Mariana Islands" OR Oman OR Mayotte OR Muscat OR Pakistan OR Palau OR Palestine OR Panama OR Paraguay OR Peru OR Philippines OR Philipines OR Phillipines OR Phillippines OR Polynesia OR "Puerto Rico" OR Qatar OR Reunion OR Rwanda OR Ruanda OR "Saint Kitts" OR "St Kitts" OR Nevis OR "Saint Lucia" OR "St Lucia" OR "Saint Vincent" OR "St Vincent" OR Grenadines OR Samoa OR "Samoan Islands" OR "Navigator Island" OR "Navigator Islands" OR "Sao Tome" OR "Saudi Arabia" OR Senegal OR | 5,882,128 |

| Search | Query | Number of Results |
| --- | --- | --- |
| | Serbia OR Montenegro OR Seychelles OR "Sierra Leone" OR Singapore OR "Sri Lanka" OR Ceylon OR "Solomon Islands" OR Somalia OR "South Africa" OR Sudan OR Suriname OR Surinam OR Swaziland OR Syria OR Tajikistan OR Tadzhikistan OR Tadjikistan OR Tadzhik OR Tanzania OR Thailand OR Togo OR "Togolese Republic" OR Tokelau OR Tonga OR Trinidad OR Tobago OR Tunisia OR Turkey OR Turkmenistan OR Turkmen OR "Turks Caicos" OR "Turks and Caicos" OR Tuvalu OR Uganda OR "United Arab Emirates" OR Uruguay OR Uzbekistan OR Uzbek OR Vanuatu OR "New Hebrides" OR Venezuela OR Vietnam OR "Viet Nam" OR "Virgin Islands" OR "West Bank" OR Yemen OR Yugoslavia OR Zambia OR Zimbabwe OR "developing population*" OR "developing world" OR "less developed countr*" OR "less developed nation*" OR "less developed world" OR "lesser developed countr*" OR "lesser developed nation*" OR "lesser developed world" OR "under developed countr*" OR "under developed nation*" OR "under developed world" OR "underdeveloped countr*" OR "underdeveloped nation*" OR "underdeveloped world" OR "middle income countr*" OR "middle income nation*" OR "middle income population*" OR "low income countr*" OR "low income nation*" OR "low income population*" OR "lower income countr*" OR "lower income nation*" OR "lower income population*" OR "underserved countr*" OR "underserved nation*" OR "underserved population*" OR "under served population*" OR "under served nation*" OR "under served population*" OR "deprived countr*" OR "deprived population*" OR "high burden countr*" OR "high burden nation*" OR "countdown countr*" OR "countdown nation*" OR "poor countr*" OR "poor nation*" OR "poor population*" OR "poor world" OR "poorer countr*" OR "poorer nation*" OR "poorer population*" OR "poorer world" OR "developing econom*" OR "less developed econom*" OR "underdeveloped econom*" OR "under developed econom*" OR "middle income econom*" OR "low income econom*" OR "lower income econom*" OR "low gdp" OR "low gnp" OR "low gross domestic" OR "low gross national" OR "lower gdp" OR "lower gnp" OR "lower gross domestic" OR "lower gross national" OR "lmic" OR "lmics" OR "third world" OR "lami countr*" OR "transitional countr*" OR "emerging econom*" OR "emerging nation*") | |
| 4 | #1 AND #2 AND #3 | 31,949 |
| 5 | #4 AND ( LIMIT-TO ( PUBYEAR , 2008 ) OR LIMIT-TO ( PUBYEAR , 2009 ) OR LIMIT-TO ( PUBYEAR , 2010 ) OR LIMIT-TO ( PUBYEAR , 2011 ) OR LIMIT-TO ( PUBYEAR , 2012 ) OR LIMIT-TO ( PUBYEAR , 2013 ) OR LIMIT-TO ( PUBYEAR , 2014 ) OR LIMIT-TO ( PUBYEAR , 2015 ) OR LIMIT-TO ( PUBYEAR , 2016 ) OR LIMIT-TO ( PUBYEAR , 2017 ) OR LIMIT-TO ( PUBYEAR , 2018 ) OR LIMIT-TO ( PUBYEAR , 2019 ) OR LIMIT-TO ( PUBYEAR , 2020 ) OR LIMIT-TO ( PUBYEAR , 2021 ) OR LIMIT-TO ( PUBYEAR , 2022 ) OR LIMIT-TO ( PUBYEAR , 2023 ) OR LIMIT-TO ( PUBYEAR , 2024 ) OR LIMIT-TO ( PUBYEAR , 2025 ) ) | 26,884 |
| 6 | #5 AND ( EXCLUDE ( DOCTYPE , "cp" ) OR EXCLUDE ( DOCTYPE , "cr" ) ) | 25,896 |

| Search | Query | Number of Results |
|---|---|---|
| 7 | #6 AND ( LIMIT-TO ( LANGUAGE , "English" ) ) | 24,038 |
| **Total** | | **24,034*** |

*Scopus displayed 24,038 but 4 were duplicates resulting in 24,034 exporting.

***Supplementary table 3: Global Health (EBSCOhost): January 8, 2025***

| Search | Query | Number of Results |
|---|---|---|
| 1 | (TI "standard precaution" OR AB "standard precaution") OR (TI "standard precautions" OR AB "standard precautions") OR (TI "infection control" OR AB "infection control") OR (TI "infection prevention" OR AB "infection prevention") OR (TI IPC OR AB IPC) OR (TI "personal protective equipment" OR AB "personal protective equipment") OR (TI PPE OR AB PPE) OR (TI "splash protection" OR AB "splash protection") OR (TI "respiratory protection" OR AB "respiratory protection") OR (TI mask OR AB mask) OR (TI masks OR AB masks) OR (TI glove OR AB glove) OR (TI gloves OR AB gloves) OR (TI gown OR AB gown) OR (TI gowns OR AB gowns) OR (TI goggles OR AB goggles) OR (TI "eye protection" OR AB "eye protection") OR (TI "face shield" OR AB "face shield") OR (TI "face shields" OR AB "face shields") OR (TI respirator* OR AB respirator*) OR (TI water OR AB water) OR (TI sanitation OR AB sanitation) OR (TI sanitary OR AB sanitary) OR (TI plumbing OR AB plumbing) OR (TI sewage OR AB sewage) OR (TI sewer* OR AB sewer*) OR (TI latrine OR AB latrine) OR (TI latrines OR AB latrines) OR (TI toilet OR AB toilet) OR (TI toilets OR AB toilets) OR (TI hygiene OR AB hygiene) OR (TI hygienic OR AB hygienic) OR (TI shower OR AB shower) OR (TI showers OR AB showers) OR (TI soap OR AB soap) OR (TI soaps OR AB soaps) OR (TI detergent OR AB detergent) OR (TI detergents OR AB detergents) OR (TI handwashing OR AB handwashing) OR (TI "hand washing" OR AB "hand washing") OR (TI "hand hygiene" OR AB "hand hygiene") OR (TI laundry OR AB laundry) OR (TI bedsheet OR AB bedsheet) OR (TI bedsheets OR AB bedsheets) OR (TI bedding OR AB bedding) OR (TI linen OR AB linen) OR (TI linens OR AB linens) OR (TI waste OR AB waste) OR (TI wastes OR AB wastes) OR (TI landfill OR AB landfill) OR (TI landfills OR AB landfills) OR (TI dump OR AB dump) OR (TI dumps OR AB dumps) OR (TI drainage OR AB drainage) OR (TI wastewater OR AB wastewater) OR (TI "waste water" OR AB "waste water") OR (TI disposal OR AB disposal) OR (TI disinfect* OR AB disinfect*) OR (TI cleaner OR AB cleaner) OR (TI cleaners OR AB cleaners) OR (TI cleaning OR AB cleaning) OR (TI cleanliness OR AB cleanliness) OR (TI janitor* OR AB janitor*) OR (TI housekeep* OR AB housekeep*) | 622,919 |
| 2 | (TI healthcare OR AB healthcare) OR (TI "health care" OR AB "health care") OR (TI hospital OR AB hospital) OR (TI hospitals OR AB hospitals) OR (TI clinic OR AB clinic) OR (TI clinics OR AB clinics) OR (TI "health facility" OR AB "health facility") OR (TI "health facilities" OR AB "health facilities") OR (TI "health center" OR AB "health center") OR (TI "health centers" OR AB | 567,774 |

| Search | Query | Number of Results |
|---|---|---|
| | "health centers") OR (TI healthcenter OR AB healthcenter) OR (TI healthcenters OR AB healthcenters) OR (TI healthcentre OR AB healthcentre) OR (TI healthcentre OR AB healthcentre) OR (TI "health centre" OR AB "health centre") OR (TI "health centres" OR AB "health centres") OR (TI "health post" OR AB "health post") OR (TI "health post" OR AB "health post") OR (TI healthpost OR AB healthpost) OR (TI healthposts OR AB healthposts) OR (TI "health setting" OR AB "health setting") OR (TI "health settings" OR AB "health settings") OR (TI "medical facility" OR AB "medical facility") OR (TI "medical facilities" OR AB "medical facilities") OR (TI "medical center" OR AB "medical center") OR (TI "medical centers" OR AB "medical centers") OR (TI "medical centre" OR AB "medical centre") OR (TI "medical centres" OR AB "medical centres") OR (TI "medical post" OR AB "medical post") OR (TI "medical posts" OR AB "medical posts") OR (TI "medical setting" OR AB "medical setting") OR (TI "medical settings" OR AB "medical settings") OR (TI "delivery facility" OR AB "delivery facility") OR (TI "delivery facilities" OR AB "delivery facilities") OR (TI "delivery center" OR AB "delivery center") OR (TI "delivery centers" OR AB "delivery centers") OR (TI "delivery centre" OR AB "delivery centre") OR (TI "delivery centres" OR AB "delivery centres") OR (TI "delivery clinic" OR AB "delivery clinic") OR (TI "delivery clinics" OR AB "delivery clinics") OR (TI "birth facility" OR AB "birth facility") OR (TI "birth facilities" OR AB "birth facilities") OR (TI "birth center" OR AB "birth center") OR (TI "birth centers" OR AB "birth centers") OR (TI "birth centre" OR AB "birth centre") OR (TI "birth centres" OR AB "birth centres") OR (TI "birth clinic" OR AB "birth clinic") OR (TI "birth clinics" OR AB "birth clinics") OR (TI "maternal facility" OR AB "maternal facility") OR (TI "maternity facility" OR AB "maternity facility") OR (TI "maternal facilities" OR AB "maternal facilities") OR (TI "maternity facilities" OR AB "maternity facilities") OR (TI "maternal center" OR AB "maternal center") OR (TI "maternity center" OR AB "maternity center") OR (TI "maternal centers" OR AB "maternal centers") OR (TI "maternity centers" OR AB "maternity centers") OR (TI "maternal centre" OR AB "maternal centre") OR (TI "maternal centers" OR AB "maternal centers") OR (TI "maternity clinic" OR AB "maternity clinic") OR (TI "maternity clinics" OR AB "maternity clinics") OR (TI "maternal clinic" OR AB "maternal clinic") OR (TI "maternal clinics" OR AB "maternal clinics") OR (TI "maternity home" OR AB "maternity home") OR (TI "maternity homes" OR AB "maternity homes") OR (TI "maternity waiting homes" OR AB "maternity waiting homes") OR (TI dispensary OR AB dispensary) OR (TI dispensaries OR AB dispensaries) | |
| 3 | (TI Afghanistan OR AB Afghanistan) OR (TI Algeria OR AB Algeria) OR (TI Angola OR AB Angola) OR (TI Anguilla OR AB Anguilla) OR (TI Antigua OR AB Antigua) OR (TI Barbuda OR AB Barbuda) OR (TI Argentina OR AB Argentina) OR (TI Armenia OR AB Armenia) OR (TI Armenian OR AB Armenian) OR (TI Aruba OR AB Aruba) OR (TI Azerbaijan OR AB Azerbaijan) OR (TI Bahamas OR AB Bahamas) OR (TI Bahrain OR AB Bahrain) OR (TI Bangladesh OR AB Bangladesh) OR (TI Barbados OR AB Barbados) OR (TI Benin OR AB Benin) | 936,763 |

| Search | Query | Number of Results |
| --- | --- | --- |
| | OR (TI Byelarus OR AB Byelarus) OR (TI Byelorussian OR AB Byelorussian) OR (TI Belarus OR AB Belarus) OR (TI Belorussian OR AB Belorussian) OR (TI Belorussia OR AB Belorussia) OR (TI Belize OR AB Belize) OR (TI Bhutan OR AB Bhutan) OR (TI Bolivia OR AB Bolivia) OR (TI Botswana OR AB Botswana) OR (TI Brazil OR AB Brazil) OR (TI Brunei OR AB Brunei) OR (TI "Burkina Faso" OR AB "Burkina Faso") OR (TI "Burkina Fasso" OR AB "Burkina Fasso") OR (TI "Upper Volta" OR AB "Upper Volta") OR (TI Burundi OR AB Burundi) OR (TI Urundi OR AB Urundi) OR (TI Cambodia OR AB Cambodia) OR (TI "Khmer Republic" OR AB "Khmer Republic") OR (TI Kampuchea OR AB Kampuchea) OR (TI Cameroon OR AB Cameroon) OR (TI Cameroons OR AB Cameroons) OR (TI Cameron OR AB Cameron) OR (TI Camerons OR AB Camerons) OR (TI "Cape Verde" OR AB "Cape Verde") OR (TI "Cayman Islands" OR AB "Cayman Islands") OR (TI "Central African Republic" OR AB "Central African Republic") OR (TI Chad OR AB Chad) OR (TI Chile OR AB Chile) OR (TI China OR AB China) OR (TI Colombia OR AB Colombia) OR (TI Comoros OR AB Comoros) OR (TI "Comoro Islands" OR AB "Comoro Islands") OR (TI Comores OR AB Comores) OR (TI Mayotte OR AB Mayotte) OR (TI Congo OR AB Congo) OR (TI Zaire OR AB Zaire) OR (TI "Cook Islands" OR AB "Cook Islands") OR (TI "Costa Rica" OR AB "Costa Rica") OR (TI "Cote d'Ivoire" OR AB "Cote d'Ivoire") OR (TI "Ivory Coast" OR AB "Ivory Coast") OR (TI Croatia OR AB Croatia) OR (TI Cuba OR AB Cuba) OR (TI Cyprus OR AB Cyprus) OR (TI Djibouti OR AB Djibouti) OR (TI "French Somaliland" OR AB "French Somaliland") OR (TI Dominica OR AB Dominica) OR (TI "Dominican Republic" OR AB "Dominican Republic") OR (TI "East Timor" OR AB "East Timor") OR (TI "East Timur" OR AB "East Timur") OR (TI "Timor Leste" OR AB "Timor Leste") OR (TI Ecuador OR AB Ecuador) OR (TI Egypt OR AB Egypt) OR (TI "United Arab Republic" OR AB "United Arab Republic") OR (TI "El Salvador" OR AB "El Salvador") OR (TI Eritrea OR AB Eritrea) OR (TI Ethiopia OR AB Ethiopia) OR (TI "Falkland Islands" OR AB "Falkland Islands") OR (TI "Las Malvinas" OR AB "Las Malvinas") OR (TI Fiji OR AB Fiji) OR (TI Gabon OR AB Gabon) OR (TI "Gabonese Republic" OR AB "Gabonese Republic") OR (TI Gambia OR AB Gambia) OR (TI Gaza OR AB Gaza) OR (TI "Georgia Republic" OR AB "Georgia Republic") OR (TI "Georgian Republic" OR AB "Georgian Republic") OR (TI Ghana OR AB Ghana) OR (TI "Gold Coast" OR AB "Gold Coast") OR (TI Greece OR AB Greece) OR (TI Grenada OR AB Grenada) OR (TI Guatemala OR AB Guatemala) OR (TI Guinea OR AB Guinea) OR (TI Guam OR AB Guam) OR (TI Guadeloupe OR AB Guadeloupe) OR (TI Guiana OR AB Guiana) OR (TI Guyana OR AB Guyana) OR (TI Haiti OR AB Haiti) OR (TI Honduras OR AB Honduras) OR (TI "Hong Kong" OR AB "Hong Kong") OR (TI India OR AB India) OR (TI Maldives OR AB Maldives) OR (TI Indonesia OR AB Indonesia) OR (TI Iran OR AB Iran) OR (TI Iraq OR AB Iraq) OR (TI Jamaica OR AB Jamaica) OR (TI Jordan OR AB Jordan) OR (TI Kazakhstan OR AB Kazakhstan) OR (TI Kazakh OR AB Kazakh) OR (TI Kenya OR AB Kenya) OR (TI Kiribati OR AB Kiribati) OR (TI Korea OR AB Korea) OR (TI Kosovo OR AB Kosovo) OR (TI Kuwait OR AB Kuwait) OR (TI Kyrgyzstan OR AB Kyrgyzstan) | |

| Search | Query | Number of Results |
|---|---|---|
| | OR (TI Kirghizia OR AB Kirghizia) OR (TI "Kyrgyz Republic" OR AB "Kyrgyz Republic") OR (TI Kirghiz OR AB Kirghiz) OR (TI Kirgizstan OR AB Kirgizstan) OR (TI "Lao PDR" OR AB "Lao PDR") OR (TI Laos OR AB Laos) OR (TI Lebanon OR AB Lebanon) OR (TI Lesotho OR AB Lesotho) OR (TI Basutoland OR AB Basutoland) OR (TI Liberia OR AB Liberia) OR (TI Libya OR AB Libya) OR (TI Macau OR AB Macau) OR (TI Madagascar OR AB Madagascar) OR (TI "Malagasy Republic" OR AB "Malagasy Republic") OR (TI Maldives OR AB Maldives) OR (TI Malaysia OR AB Malaysia) OR (TI Malaya OR AB Malaya) OR (TI Malay OR AB Malay) OR (TI Sabah OR AB Sabah) OR (TI Sarawak OR AB Sarawak) OR (TI Malawi OR AB Malawi) OR (TI Nyasaland OR AB Nyasaland) OR (TI Mali OR AB Mali) OR (TI Malta OR AB Malta) OR (TI "Marshall Islands" OR AB "Marshall Islands") OR (TI Martinique OR AB Martinique) OR (TI Mauritania OR AB Mauritania) OR (TI Mauritius OR AB Mauritius) OR (TI "Agalega Islands" OR AB "Agalega Islands") OR (TI Mexico OR AB Mexico) OR (TI Micronesia OR AB Micronesia) OR (TI "Middle East" OR AB "Middle East") OR (TI Mongolia OR AB Mongolia) OR (TI Montserrat OR AB Montserrat) OR (TI Morocco OR AB Morocco) OR (TI Ifni OR AB Ifni) OR (TI Mozambique OR AB Mozambique) OR (TI Myanmar OR AB Myanmar) OR (TI Myanma OR AB Myanma) OR (TI Burma OR AB Burma) OR (TI Namibia OR AB Namibia) OR (TI Nauru OR AB Nauru) OR (TI Nepal OR AB Nepal) OR (TI Niui OR AB Niui) OR (TI "Netherlands Antilles" OR AB "Netherlands Antilles") OR (TI "New Caledonia" OR AB "New Caledonia") OR (TI Nicaragua OR AB Nicaragua) OR (TI Niger OR AB Niger) OR (TI Nigeria OR AB Nigeria) OR (TI "Northern Mariana Islands" OR AB "Northern Mariana Islands") OR (TI Oman OR AB Oman) OR (TI Mayotte OR AB Mayotte) OR (TI Muscat OR AB Muscat) OR (TI Pakistan OR AB Pakistan) OR (TI Palau OR AB Palau) OR (TI Palestine OR AB Palestine) OR (TI Panama OR AB Panama) OR (TI Paraguay OR AB Paraguay) OR (TI Peru OR AB Peru) OR (TI Philippines OR AB Philippines) OR (TI Philipines OR AB Philipines) OR (TI Phillipines OR AB Phillipines) OR (TI Phillippines OR AB Phillippines) OR (TI Polynesia OR AB Polynesia) OR (TI "Puerto Rico" OR AB "Puerto Rico") OR (TI Qatar OR AB Qatar) OR (TI Reunion OR AB Reunion) OR (TI Rwanda OR AB Rwanda) OR (TI Ruanda OR AB Ruanda) OR (TI "Saint Kitts" OR AB "Saint Kitts") OR (TI "St Kitts" OR AB "St Kitts") OR (TI Nevis OR AB Nevis) OR (TI "Saint Lucia" OR AB "Saint Lucia") OR (TI "St Lucia" OR AB "St Lucia") OR (TI "Saint Vincent" OR AB "Saint Vincent") OR (TI "St Vincent" OR AB "St Vincent") OR (TI Grenadines OR AB Grenadines) OR (TI Samoa OR AB Samoa) OR (TI "Samoan Islands" OR AB "Samoan Islands") OR (TI "Navigator Island" OR AB "Navigator Island") OR (TI "Navigator Islands" OR AB "Navigator Islands") OR (TI "Sao Tome" OR AB "Sao Tome") OR (TI "Saudi Arabia" OR AB "Saudi Arabia") OR (TI Senegal OR AB Senegal) OR (TI Serbia OR AB Serbia) OR (TI Montenegro OR AB Montenegro) OR (TI Seychelles OR AB Seychelles) OR (TI "Sierra Leone" OR AB "Sierra Leone") OR (TI Singapore OR AB Singapore) OR (TI "Sri Lanka" OR AB "Sri Lanka") OR (TI Ceylon OR AB Ceylon) OR (TI "Solomon Islands" OR AB "Solomon Islands") OR (TI Somalia OR AB Somalia) OR (TI | |

| Search | Query | Number of Results |
|---|---|---|
| | "South Africa" OR AB "South Africa") OR (TI Sudan OR AB Sudan) OR (TI Suriname OR AB Suriname) OR (TI Surinam OR AB Surinam) OR (TI Swaziland OR AB Swaziland) OR (TI Syria OR AB Syria) OR (TI Tajikistan OR AB Tajikistan) OR (TI Tadzhikistan OR AB Tadzhikistan) OR (TI Tadjikistan OR AB Tadjikistan) OR (TI Tadzhik OR AB Tadzhik) OR (TI Tanzania OR AB Tanzania) OR (TI Thailand OR AB Thailand) OR (TI Togo OR AB Togo) OR (TI "Togolese Republic" OR AB "Togolese Republic") OR (TI Tokelau OR AB Tokelau) OR (TI Tonga OR AB Tonga) OR (TI Trinidad OR AB Trinidad) OR (TI Tobago OR AB Tobago) OR (TI Tunisia OR AB Tunisia) OR (TI Turkey OR AB Turkey) OR (TI Turkmenistan OR AB Turkmenistan) OR (TI Turkmen OR AB Turkmen) OR (TI "Turks Caicos" OR AB "Turks Caicos") OR (TI "Turks and Caicos" OR AB "Turks and Caicos") OR (TI Tuvalu OR AB Tuvalu) OR (TI Uganda OR AB Uganda) OR (TI "United Arab Emirates" OR AB "United Arab Emirates") OR (TI Uruguay OR AB Uruguay) OR (TI Uzbekistan OR AB Uzbekistan) OR (TI Uzbek OR AB Uzbek) OR (TI Vanuatu OR AB Vanuatu) OR (TI "New Hebrides" OR AB "New Hebrides") OR (TI Venezuela OR AB Venezuela) OR (TI Vietnam OR AB Vietnam) OR (TI "Viet Nam" OR AB "Viet Nam") OR (TI "Virgin Islands" OR AB "Virgin Islands") OR (TI "West Bank" OR AB "West Bank") OR (TI Yemen OR AB Yemen) OR (TI Yugoslavia OR AB Yugoslavia) OR (TI Zambia OR AB Zambia) OR (TI Zimbabwe OR AB Zimbabwe) OR TI (“developing population*” OR “developing world” OR “less developed countr*” OR “less developed nation*” OR “less developed world” OR “lesser developed countr*” OR “lesser developed nation*” OR “lesser developed world” OR “under developed countr*” OR “under developed nation*” OR “under developed world” OR “underdeveloped countr*” OR “underdeveloped nation*” OR “underdeveloped world” OR “middle income countr*” OR “middle income nation*” OR “middle income population*” OR “low income countr*” OR “low income nation*” OR “low income population*” OR “lower income countr*” OR “lower income nation*” OR “lower income population*” OR “underserved countr*” OR “underserved nation*” OR “underserved population*” OR “under served population*” OR “under served nation*” OR “under served population*” OR “deprived countr*” OR “deprived population*” OR “high burden countr*” OR “high burden nation*” OR “countdown countr*” OR “countdown nation*” OR “poor countr*” OR “poor nation*” OR “poor population*” OR “poor world” OR “poorer countr*” OR “poorer nation*” OR “poorer population*” OR “poorer world” OR “developing econom*” OR “less developed econom*” OR “underdeveloped econom*” OR “under developed econom*” OR “middle income econom*” OR “low income econom*” OR “lower income econom*” OR “low gdp” OR “low gnp” OR “low gross domestic” OR “low gross national” OR “lower gdp” OR “lower gnp” OR “lower gross domestic” OR “lower gross national” OR “lmic” OR “lmics” OR “third world” OR “lami countr*” OR “transitional countr*” OR “emerging econom*” OR “emerging nation*”) OR AB (“developing population*” OR “developing world” OR “less developed countr*” OR “less developed nation*” OR “less | |

| Search | Query | Number of Results |
|---|---|---|
| | developed world" OR "lesser developed countr*" OR "lesser developed nation*" OR "lesser developed world" OR "under developed countr*" OR "under developed nation*" OR "under developed world" OR "underdeveloped countr*" OR "underdeveloped nation*" OR "underdeveloped world" OR "middle income countr*" OR "middle income nation*" OR "middle income population*" OR "low income countr*" OR "low income nation*" OR "low income population*" OR "lower income countr*" OR "lower income nation*" OR "lower income population*" OR "underserved countr*" OR "underserved nation*" OR "underserved population*" OR "under served population*" OR "under served nation*" OR "under served population*" OR "deprived countr*" OR "deprived population*" OR "high burden countr*" OR "high burden nation*" OR "countdown countr*" OR "countdown nation*" OR "poor countr*" OR "poor nation*" OR "poor population*" OR "poor world" OR "poorer countr*" OR "poorer nation*" OR "poorer population*" OR "poorer world" OR "developing econom*" OR "less developed econom*" OR "underdeveloped econom*" OR "under developed econom*" OR "middle income econom*" OR "low income econom*" OR "lower income econom*" OR "low gdp" OR "low gnp" OR "low gross domestic" OR "low gross national" OR "lower gdp" OR "lower gnp" OR "lower gross domestic" OR "lower gross national" OR "lmic" OR "lmics" OR "third world" OR "lami countr*" OR "transitional countr*" OR "emerging econom*" OR "emerging nation*") | |
| 4 | #1 AND #2 AND #3 | 27,732 |
| 5 | #4 AND Limit by: Publication Year: 20080101-20250108 | 22,985 |
| 6 | #5 AND Limit by: Source Type: Academic Journals OR Reports | 22,336 |
| 7 | #6 AND Narrow by: English Language | 20,437 |
| **Total** | | **20,437** |

**Tag categories for evidence map of environmental health services in healthcare facilities**

Supplementary Table 4. Tag categories and definitions for the classification of studies included in the evidence map of environmental health services in healthcare settings in low- and middle-income countries

| Environmental health services | |
|---|---|
| Tag | Definition |
| Water | - Water infrastructure<br>- Water quality<br>- Water quantity/accessibility<br>- Behaviors related to water testing and treatment |
| Sanitation | - Toilet or latrines<br>- Menstrual hygiene management<br>- Wastewater treatment and disposal |
| Hygiene at the point of care | - Healthcare worker hand hygiene behavior/compliance – including studies that measure knowledge or observe behaviors of hygiene during clinical care<br>- Studies that apply the WHO Five Moments for Hand Hygiene guidelines<br>- Studies that examine hand hygiene specifically for infection prevention and control<br>- Studies that examine "universal precautions" or "standard precautions" for infection control if services are not otherwise specified<br>- Hand hygiene supplies available in patient care settings/at the point of care (e.g., soap, alcohol-based hand rub, surgical scrub)<br>- Scrubbing, eye wash stations, and other infrastructure/supplies for body decontamination associated with clinical care<br>- Usage of personal protective equipment (PPE) during patient care<br>- Microbiology studies of hand contamination of healthcare workers |
| Hygiene outside the point of care | - Hand hygiene supplies and behaviors in settings other than the point of care (e.g., waiting rooms, offices, laboratories, bathrooms)<br>- Hand hygiene behaviors by patients and caregivers<br>- IPC measures performed outside the point of care – e.g., sterilization and disinfection of items that are used in a patient care environment<br>- Use of PPE outside the point of care, except for waste management and cleaning<br>- Shower and bathing facilities for patients |
| Waste | - Healthcare waste, including sharps safety, infectious waste, hazardous waste, and general waste<br>- Waste collection, segregation, storage, treatment, and disposal<br>- Infrastructure for waste management (e.g., incinerators)<br>- Use of personal protective equipment (PPE) during waste handling and disposal |
| Cleaning | - Supplies, behaviors, and infrastructure for general cleaning of surfaces in patient and non-patient spaces, cleaning of infectious or hazardous spills, or disinfecting surfaces and devices<br>- Cleaning supplies (e.g., brooms, mops, disinfectants, hypochlorite)<br>- Studies reporting levels of cleanliness on bathrooms, wards, etc. (if bathrooms, also tag as sanitation) |

| | |
|---|---|
| | - Studies that take samples from surfaces or medical devices to assess microbiological contamination and cleanliness |
| Laundry | Infrastructure, supplies, and behaviors related to laundry and linens |
| Study design | |
| Tag | Definition |
| Qualitative | Uses qualitative methods, including interviews, focus groups, workshops, ethnographic research, and expert elicitation. |
| Review | A systematic, scoping, critical, etc. review of published literature that does not include primary data collection. |
| Cross-sectional survey | - Uses surveys, structured observations, environmental sample collection, or another quantitative method to examine environmental health services at *one point*. It also includes simulation/modeling studies based on survey data collection.<br>- Does not evaluate an intervention/compare an intervention to a control |
| Longitudinal survey | - Uses surveys, structured observations, environmental sample collection, or other quantitative methods to examine services repeatedly across multiple (two or more) time points.<br>- Does not evaluate an intervention |
| Non-experimental intervention | Measures the effect of an intervention but does *not* use randomized controlled trial methods (in other words, participants are not randomly assigned to the intervention or control group). Includes before-and-after studies, interrupted time series, case-control, and natural experiments. |
| Experimental intervention | A randomized controlled trial/experiment to measure the effect of an intervention. One group receives an intervention, while the other group serves as a control. Participants in the intervention and control groups are selected randomly from the population. |
| Other/not specified method | The study is a research study, but it uses a method that is not covered in the above definitions. |
| Policy/practice relevance: Eight Practical Steps | |
| Tag | Definition |
| Step 1A: Conduct situation analysis | - Assessments of the policy landscape or “enabling environment” for environmental health services<br>- Studies that describe management-level, government, or policy-level readiness or barriers for WASH improvements (from the level of the facility up to the national level) |
| Step 1B: Conduct baseline assessment | - Quantitative descriptions of environmental coverage/access<br>- Studies that use an assessment tool to evaluate baseline conditions (e.g., the WASH FIT tool)<br>- Studies that describe healthcare worker or patient knowledge, attitudes, practices, and behaviors (KAP) related to environmental health services |
| Step 1C: Formative research | - Studies that propose “formative pathways” for environmental health services, i.e., examine links between exposures and outcomes along the impact pathway<br>- Uses any method to establish associations between variables – e.g., regression, observational, qualitative research<br>- Does not include an intervention |

| | |
|---|---|
| | - Studies that examine "associations," "predictors," or use regression analysis are likely included here. Qualitative studies that describe worker or patient perspectives on environmental health services are likely included here. |
| Step 2: Set targets and establish a coordination mechanism | - Studies that report on the creation of national plans, roadmaps, and task forces for environmental health services<br>- Budgeting and costing for environmental health services |
| Step 3: Establish national WASH standards or healthcare waste standards | - Studies that describe the process of developing national standards for environmental health services or infection control<br>- Studies that examine standards and accountability mechanisms for environmental health services |
| Step 4: Improve and maintain environmental health services | - Studies that (a) modify environmental health services through an intervention, program, or implementation strategy and (b) study the effects of these modifications<br>- Any intervention – including interventions to modify environmental infrastructure, behaviors, operations/maintenance, etc.<br>- Any study design, including experimental, quasi-experimental, or pretest-posttest<br>- Impact and process evaluations for interventions or policies<br>- Studies that evaluate factors influencing long-term operations, maintenance, and sustainability of interventions/programs |
| Step 5: Monitor and review data | A longitudinal assessment of environmental health services that is conducted with routine monitoring data (e.g., district health information system data) |
| Step 6: Develop a health workforce | - Studies to assess healthcare worker knowledge, attitudes, and behaviors related to environmental health services<br>- Studies that involve quality improvement or training to improve healthcare worker knowledge/behavior<br>- Includes studies that assess or improve healthcare worker knowledge and behavior related to hand hygiene, infection control measures, waste management, cleaning, etc.<br>- Studies of medical student knowledge/behaviors |
| Step 7: Engage communities | Uses participatory research approaches to engage patients or other communities outside of the healthcare system |
| Topic/population focus | |
| Tag | Definition |
| COVID | Examines environmental health services and related behaviors in healthcare facilities during the COVID-19 pandemic, COVID-19 preparedness, or impacts of COVID-19 |
| Other epidemics | Examines environmental health services and related behaviors in healthcare facilities during non-COVID outbreaks, epidemics, and pandemics. E.g., Ebola, Lassa fever, influenza |
| Climate | Evaluates relationships between climate and healthcare facility environmental health services – including topics such as climate resilience, impacts of extreme weather events, and climate emergency preparedness |
| Maternal | Studies that examine environmental health services in maternal, child, and newborn health settings.<br>Includes studies conducted in healthcare settings where the primary purpose is maternal and child health (including antenatal care, labor and delivery, postnatal/neonatal, and pediatrics). This could include facilities specializing in maternal/child health or specialized wards/departments within a more general facility. |

| | |
|---|---|
| | Includes studies that examine the impacts of environmental health services among populations of pregnant women, newborns, or children under age five |
